# Unraveling the Single-Cell Spatial Landscapes of Melanoma Brain Metastases

**DOI:** 10.1101/2025.08.29.25334740

**Authors:** Artür Manukyan, Kristin Peters, Helena Radbruch, Federico Fusco, Felicitas Geserick, Florian Rossner, Annika Lehmann, Karsten Kleo, Thomas Conrad, Janine Altmüller, Izabela Plumbom, Altuna Akalin, Karsten Jürchott, Emanuel Wyler, Markus Landthaler, Claire Delbridge, Matthias Sendler, Josefine Radke, Torben Redmer

## Abstract

The cellular composition of the tumor ecosystem critically influences responses to immune checkpoint inhibitors (ICi) and BRAF inhibitors (BRAFi), however the specific dynamics driving the progression of melanoma brain metastases (MBMs) remain poorly understood. Here, we performed single cell-resolved spatial transcriptomics profiling of multiple tissue cores (n=47) of distinct solitary (n=12) and concordant multiple MBM (n=22), CNS tissue (n=9) and whole sections including one matched primary (n=4) of MBMs across various developmental stages and therapeutic regimens. Our approach uncovered both shared and distinct evolutionary patterns of MBM progression, highlighting pronounced cellular and spatial heterogeneity while identifying key tumor subsets defined by MET or NGFR expression. Notably, spatial profiling of tumors progressing under ICi or BRAFi therapy identified a population of BZW2^+^ tumor cells that promotes an immunosuppressive microenvironment, thereby impeding immune cell infiltration and intratumoral dispersion. Elevated BZW2 expression was associated with ICi resistance and inversely correlated with the antigen transporter TAP1 and the activation of interferon/STAT1 signaling. Spatially aware automated profiling of immune-enriched niches distinguished “hot” niches - characterized by tumor expression of TAP1, PD-L2 and HLA-DRA - from “cold” zones, which were characterized by high levels of BZW2 and the transcriptional regulator SOX4. This translational suppressor co-localized with the clinically targetable MET receptor in a subset of tumor cells exhibiting distinct immune evasion features.

In summary, we provide insights into the single-cell and spatially resolved landscapes of MBM, identifying BZW2/SOX4 and TAP1 as potential determinants of ICi response.

## Introduction

Tumor development and progression are highly dynamic processes^1^ that are temporally defined and driven by a complex interplay of genetic aberrations^2^, environmental factors, and spatial mechanisms which in turn are regulated by interactions with the tumor microenvironment (TME). While the genetic landscape of tumors provides a fundamental blueprint for their progression^3^, the emergence of subclonal populations^4^ and the crosstalk between tumor and stromal cells, particularly involving soluble factors and extracellular vesicles^5^ play crucial roles in shaping the heterogeneity and therapeutic resistance observed in cancer.

The development of brain metastases is observed in 40–60% of melanoma patients during the course of the disease and, as in patients with lung and breast cancer^6^ represents a major contributor to disease progression and poor prognosis^7–9^. Thorough spatial profiling of breast cancer and lung cancer-related brain metastases identified several different cell types, sub classes and molecular programs^10^, however single cell-resolved spatial characterizations of MBM remain elusive. In the context of brain metastases, astrocytes, oligodendrocytes and tumor-associated macrophages (TAMs) are key cell interactors and contribute to the establishment of distinct tumor niches within the brain^11^.

These cell types facilitate the maintenance and progression of metastatic tumor cells and thereby influence the transition from initially small and asymptomatic brain micrometastases to symptomatic macrometastases^11^ through intricate signaling networks^12^. Currently, therapeutic targeting of the PD-1/PD-L1 axis or LAG-3 interactions with neutralizing antibodies represents one of the most important strategies for restoring T cell activation and enabling tumor cell recognition and elimination^13^. Despite a subset of patients showing complete intracranial response (CR) to ICi, immune resistance programs (IRPs) emerge in the majority of patients and ultimately determine the clinical response of MBM to anti-PD-1 therapy^14,15^.

Likely, environmental stresses induced by BRAFi targeting the mutated and hyperactivated BRAF kinase (BRAF^V600E/K/R^), as well as by ICi aimed at blocking tumor growth, can remodel the TME and alter the dynamics of tumor-stromal interactions^16^.

These observations highlight the complexity of the TME and underscore the limitations of current therapies in overcoming tumor immune evasion and therapy resistance.

Previous studies identified gene signatures predicting the response of melanoma to ICi^14,17,18^, including a tumor-intrinsic NGFR (nerve growth factor receptor)-associated gene signature predicting anti-PD-1 therapy resistance^19^. However, signature genes likely reflect multiple concerted molecular mechanisms of immune cell (T cell) suppression, involving potentially interacting tumor- and stromal cell-associated mechanisms. The insufficient immune cell infiltration into MBM remains a major barrier to effective therapy.

Epithelial-like MBM, characterized by expression of Ecad (CDH1), adopt an NGFR^+^ cell phenotype in response to antigen-specific cytotoxic T cells^19^ or therapeutic interventions^20^. Hence, only a minority of typically early-stage or therapy-naïve tumors might exhibit high levels of immune cell infiltration. Therefore, ICi therapy might be ineffective in MBM featuring low levels of tumor immune cell infiltration and IFNγ scores^21^ as well as low PD-L1 expression^22^.

The efficacy of immune checkpoint inhibitor (ICi) therapy is likely influenced by additional, predominantly spatially regulated mechanisms, such as the presentation of tumor antigens to adjacent immune cells^23^. Moreover, the different developmental stages and molecular phenotypes of MBM, may determine the response to therapeutic interventions and drive the transition from a tumor growth-suppressive to an inflammatory and progressively evolving microenvironment that promotes tumor growth and metastasis^24^.

Brain metastases likely consist of spatial areas with varying levels of sensitivity and resistance to therapy, representing a major challenge for the development of more effective treatments. To address these challenges, we have employed spatial transcriptomic profiling to investigate the molecular landscapes of concordant multiple MBM (n=22) and solitary MBM (n=16) including whole sections of matched primary and brain metastasis (n=2) and of matched MBM (n=2) from different intracranial sites and normal CNS (n=9). Here, we present single-cell and spatially resolved heterogenic landscapes of MBM across diverse therapeutic and developmental stages.

## Methods

### Patient samples

All procedures performed in this study were in accordance with the ethical standards of the respective institutional research committees and with the 1964 Helsinki Declaration and its later amendments or comparable ethical standards. All patients gave written informed consent for the collection and scientific use of tumor material which was collected at the Department of Neuropathology, Charité-Universitätsmedizin Berlin, Germany and Department of Pathology, Universitätsmedizin Greifswald, Greifswald, Germany. The usage of archived (FFPE) melanoma and central nervous system-derived control samples has been reviewed and approved by the Ethics Committee of the Charité (EA1/107/17 and EA1/075/19) and Universitätsmedizin Greifswald (BB 001/23).

### Proteome analyses of ICI-resistant patients

The present study included proteomic data from patients with melanoma brain metastases derived from a previously published retrospective cohort of advanced melanoma cases^25^. All patients had histologically confirmed metastatic melanoma, underwent surgical resection of brain metastatic lesions and received immune checkpoint inhibitor (ICI) therapy in the metastatic setting. The analyzed subset comprised 19 patients (mean age 59.1 years, median 60 years, range 30–83 years), including 10 males and 9 females. Cutaneous melanoma was the most frequent subtype (52.6%), followed by melanoma of unknown primary (36.8%) and uveal melanoma (10.5%). BRAF mutations were detected in 52.6% of cases. Brain metastases were predominantly located in the frontal and temporal regions. Proteomic analyses were performed as previously described^25^. Tumor tissue obtained from formalin-fixed paraffin-embedded (FFPE) samples was processed using a single-pot solid-phase-enhanced sample preparation (SP3) workflow. Peptide analysis was conducted by nanoLC-FAIMS-MS/MS using an Evosep One (Evosep) liquid chromatography system coupled to an Orbitrap Exploris 480 mass spectrometer (Thermo Fisher Scientific). Protein identification and label-free quantification were performed using the FragPipe (Nesvizhskii Lab, University of Michigan, v22.0) computational platform including MSFragger and IonQuant. Full details regarding patient characteristics, sample processing, mass spectrometry acquisition parameters, and bioinformatic analysis are provided in the original publication^25^.

### Tissue microarray (TMA)

Hematoxylin and eosin (H&E)-stained brain metastasis tissue sections were examined to identify morphologically representative regions for inclusion in the tissue microarray (TMA) block. For each case, one to three tumor areas were selected under a conventional microscope (Nikon Eclipse Ni-U, Düsseldorf, Germany). Selected regions were marked on the slides and matched to the corresponding donor blocks. TMA construction was performed using a Manual Tissue Arrayer Model I (MTA-1; Estigen OÜ, Tartu, Estonia). A blank paraffin recipient block was first prepared, and cylindrical holes (1.5 mm diameter) were punched into it. Tissue cores of the same diameter were then extracted from the marked regions of the donor blocks and transferred into the recipient block, resulting in a TMA block containing 14 tissue cores (see graphical abstract, Figure 1). The completed TMA block was incubated at 60 °C for approximately 20 minutes to fuse the tissue cores with the paraffin. Finally, the block was sectioned and stained for downstream analysis.

**Figure 1:**
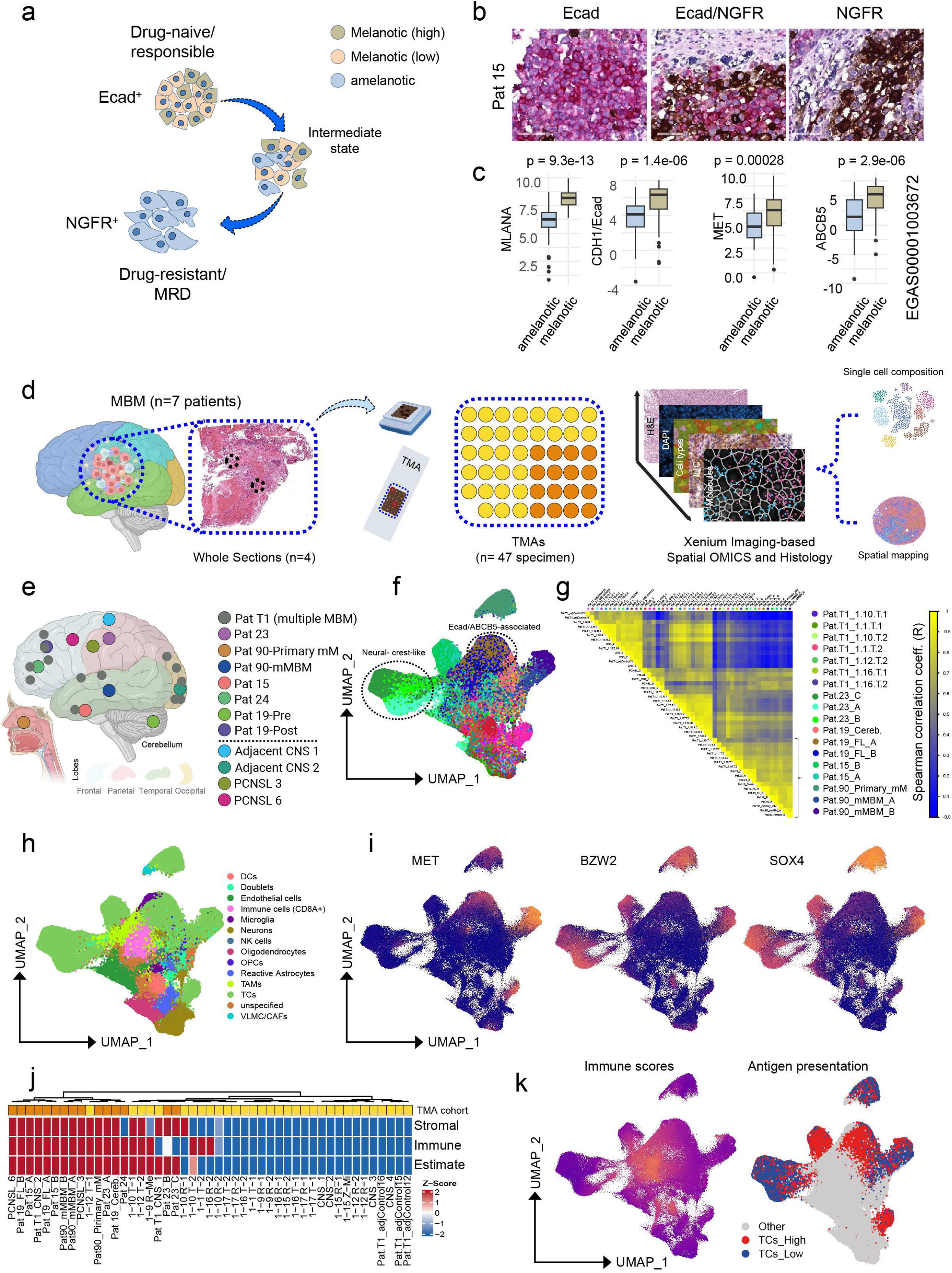
E-cadherin and NGFR define molecular subtypes of MBM. **a.)** Scheme showing the transition of putatively early stage E-cadherin (Ecad^+^) expressing MBM which transit into Ecad/NGFR intermediate and NGFR^+^ states. **b.)** Immunostaining of Ecad (red) and NGFR (brown) of MBM (Pat 15), validating the different developmental stages shown in (a). **c.)** Box plots showing expression levels of MLANA, Ecad (CDH1), MET and ABCB5 in MBM in relationship to the phenotype (amelanotic, melanotic). **d.)** Spatial transcriptomics analysis workflow illustrating tumor collection, TMA preparation, and subsequent processing and analysis of single-cell and spatial data from TMA and whole tumor slides. e.) Information about the anatomic sites of MBM. **f.)** UMAP visualization of individual cells from TMAs (n=2). Clustered samples that exhibit neural crest-like or Ecad/ABCB5-associated phenotypes are indicated. **g.)** Correlation heat map showing the clustering of samples regarding the molecular phenotype and content of CNS tissue and tumor cells (upper panel). **h.)** UMAP visualization of individual cells from samples shown in (f), showing marker-defined cell types, TCs: tumor cells; DCs: dendritic cells; NK cells: natural killer cells; OPCs: oligodendrocyte precursor cells; TAMs: tumor-associated macrophages; VLMC/CAFs: vascular leptomeningeal cells/cancer-associated fibroblasts. UMAP presentation of expression of MET (c-MET), BZW2 and SOX4 in individual cells of samples shown in (f). **j.)** Heat map showing levels (color coded) of immune, stromal and estimate scores of TMA samples. Controls and CNS indicate samples from tumor-adjacent (adj.) CNS (central nervous system) or CNS from non-cancer patients. **k.)** UMAPs showing the level of immune scores in single cells (left) and tumor cells featuring high or low expression of genes involved in antigen presentation. Non-tumor cells are indicated (Other).

### DNA extraction and quantification

DNA was extracted from FFPE tissue using the Promega® RSC DNA FFPE Kit. Samples were pretreated with 300 µL mineral oil at 80 °C for 2 minutes, followed by lysis with proteinase K for 30 minutes at 56 °C. Extraction and quantification were performed automatically using the Maxwell® RSC instrument (AS4500) and Quantus fluorometer (QuantiFluor ONE dsDNA, Promega, USA).

### Genetic Profiling of MBM using TruSight Oncology 500 (TSO500)

Library preparation followed the manufacturer’s protocol (Illumina, USA). Briefly, 100 ng DNA was sonicated using a Covaris ME220 sonicator (settings per Illumina, Table S1) to yield fragments averaging 245 bp, confirmed via TapeStation 4150 (Agilent, USA). After end-repair and A-tailing, adapters with unique molecular identifiers (UMIs) were ligated, followed by index PCR. Target enrichment involved two hybridization steps with biotin-linked probes, captured using streptavidin magnetic beads, and amplified by PCR. Libraries were bead-normalized, pooled, denatured, diluted, and sequenced on an Illumina NextSeq 550 DX (2×101 bp paired-end, dual index). Base call files (bcl) were processed using Illumina DRAGEN TruSight Oncology 500 Analysis Software v2.5.3 for FASTQ generation, alignment to hg19, variant calling, and QC evaluation (Q30, coverage >100×, contamination). All metrics met Illumina specifications. Variants from DRAGEN were further annotated via MyVariant.info’s REST API, queried with the MyVariant Python module. Variants considered included exonic SNVs, indels, splice mutations, and clinically relevant entries (ClinVar). Polymorphisms were excluded based on the 1000 Genomes project, with a mutation allele frequency threshold of 5%. Benign and likely benign variants were excluded from reporting.

### Methylome profiling Infinium MethylationEPIC v2 array

For global methylome analysis, 100-250 ng genomic DNA were subjected to bisulfite conversion using the EZ DNA Methylation-Gold Kit (Zymo Research) according to the manufacturer’s protocol. Subsequently, samples were analyzed on the Infinium MethylationEPICv2 Kit (Illumina) according to the manufacturer’s recommendations to obtain genome-wide data from 950,000 CpG positions. Raw data from Illumina Epic arrays were preprocessed and analyzed in the standard workflow of the packages RnBeads^26^ and watermelon^27^ and CNV profiles were calculated using the conumee package^28^.

### Panel design

The panel was designed as previously described. Panel genes are shown in Table S13.

### Spatial transcriptomics analysis

Xenium in situ analysis (10X Genomics) was performed with the human brain base panel (hBrain_v1, 266 genes) together with a 100 gene custom add-on panel. TMA and whole tumor sections were prepared and processed according to manufacturer’s guidelines (CG000578, Rev D; CG000760, Rev A). Briefly, the sections underwent sequential deparaffinization and permeabilization, followed by overnight hybridization with custom probes at 50°C, rolling circle amplification, and slide-read out in the Xenium Analyzer. After run completion, H&E staining was performed following Xenium guidelines (CG000613, B).

### Processing and analysis of data

Spatial analysis of the Xenium readouts of TMA and MBM samples were analyzed using the VoltRon (version 0.2.0) R package^29^. Individual TMA cores were interactively separated into distinct samples using the *subset(object, interactive = TRUE)* function. TMA cores were grouped into patient/donor of origin prior to downstream analysis. Cells with transcript counts less than 5 were removed prior to clustering. Prior to clustering and cell type annotation, post Xenium H&E images of TMA cores and remaining samples were automatically aligned to DAPI channels of the Xenium data stored in the associated VoltRon objects using the *registerSpatialData* function. Seurat (version 5) package^30^ was used to perform downstream unbiased clustering of Xenium cells from both TMA cores and the remaining MBM samples. Log normalized counts (*NormalizeData* function) was performed with a scaling factor = 100. This is followed by dimensionality reduction with PCA and UMAP, and then clustering with shared nearest neighbor graphs (*FindNeighbors* and *FindClusters* function) using parameters specific for each sample. UMAP of Xenium cells across all TMAs were calculated using Harmony-based^31^ integrated PCA embeddings of all TMAs. Marker analysis was performed to detect differentially expressed (DE) genes specific to each cell cluster (*FindAllMarkers* function). Cell types were annotated using a hybrid approach, specifically by **(i)** investigating DE genes and **(ii)** interactively visualizing clustered cells and aligned H&E images using TissUUmaps tool^32^. VoltRon objects were converted into anndata (h5ad) format prior to interactive visualization using the *as.AnnData* function. We used the AddModuleScore function from Seurat to calculate the immune module scores using a predetermined set of genes. Cells associated with hot and cold regions, which are areas of around the cells with high immune score, were automatically detected via hot spot analysis utilities available in VoltRon. We first constructed a spatial neighborhood graph to identify pairs of cells within 40 or 50 pixel distances (*getSpatialNeighbors* function), and tested significant local changes in the immune score for each cell using Getis-Ord^33^ statistics. To detect significant changes of gene expression across hot and cold regions we aggregated the count of cells annotated as hot and cold, and then performed DE analysis with the DESeq2 R package^34^. Aggregation was performed across (i) tumor cells and cells of the TME or (ii) only tumor cells depending on the genes whose expression were compared across hot and cold niches (Table S4).

### Immunohistochemistry

Automated immunohistochemical staining of formalin-fixed, paraffin-embedded (FFPE) tissue sections (2 μm thick) was performed on a VENTANA Benchmark XT automated staining instrument (Ventana Medical Systems, Tucson, AZ) according to the manufacturer’s instructions. The primary and secondary antibodies used are listed in the Methods section. Slides were deparaffinized using EZ Prep solution for 30 minutes at 75 °C. Antigen retrieval was performed on the automated stainer using CC1 solution for 60 minutes at 95 °C. Primary antibodies were then applied and visualized with either the iVIEW DAB Detection Kit or the ultraView Universal Alkaline Phosphatase Red Detection Kit (Ventana Medical Systems). All slides were counterstained with hematoxylin for 8 minutes. Immunohistochemical results were independently evaluated by two consultants in neuropathology and pathology based on predominant staining intensity. Omission of the primary antibodies, as a control for nonspecific secondary antibody binding, resulted in absence of labeling. For validation, positive control tissues - fixed and processed in the same manner as the test sections and known to contain the target molecule (e.g., colon carcinoma) - were included in each staining run.

### MET-FISH analysis

MET-FISH for evaluation of MET amplification was performed as previously described^35^.

### Multiplex staining of TMAs using the MACSima imaging system

Slides containing paraffin-embedded TMAs were used for multiplex staining. Deparaffinization was performed using xylene (10 min), methanol (5 min), 100% ethanol (3 min), 95% ethanol (3 min), 80% ethanol (3 min), 70% ethanol (3 min), 60% ethanol (3 min), and PBS (3 min). Antigen retrieval was performed using Target Retrieval Solution (Dako, Ref. S1699) in a pressure cooker for 20 minutes. The slides were placed on imaging frames (MACSwell(TM) One Small Imaging Frames, 130-126-794, Miltenyi BioTech) for the subsequent labeling cycles. The slides were blocked with FcR Blocking Reagent, human (130-059-901, Miltenyi BioTech) and pre-stained with DAPI. After DAPI staining, the slides were washed with a MACSima run buffer (130-121-565, Miltenyi). The MACSima run was performed in 5 cycles, with a bleaching step applied to remove previous labeling. The image datasets for each ROI, including all staining and autofluorescence images in TIFF format, were analyzed using MACS iQ View software (version 1.3.3). Following the MACSima run, H&E staining of the TMA slide was performed and scanned using the Sysmex Slide Scanner MIDI-II from 3D Histech.

## Results

### Molecular signatures pre-define cellular subsets of MBM

Cellular plasticity drives the distinct differentiation stages of melanoma, spanning from melanotic to amelanotic states (Figure 1a). We previously demonstrated that E-cadherin (Ecad) and NGFR govern divergent molecular programs in melanoma brain metastases (MBM) that correlate with pigmentation status^20^. Specifically, Ecad, MET (c-MET) and ABCB5 characterize the pigmented, differentiated phenotype (Figures 1b, c, S1a). In contrast, NGFR and associated genes define a de-differentiated, neural crest stem-like^36,37^ and invasive state^20^. We observed that the de-differentiated, NGFR^+^ tumors were characterized by epithelial-mesenchymal transition (EMT), TNFα/NFκB signaling, angiogenesis, and active mitosis (Figure S1b, left panel). Conversely, the Ecad^+^, melanotic cells relied on oxidative phosphorylation (OXPHOS), fatty acid metabolism, cholesterol homeostasis, and MTORC1 signaling (Figure S1b, right panel). Moreover, we observed potential association of the Ecad status with response to immune checkpoint (ICi) or BRAF inhibitor (BRAFi) therapies. Particularly, the resistance to PD-1 inhibition was likely associated with the tumor phenotypes (Figure S1c).

The pattern of infiltration of immune cells vastly determines the response of tumors to immune checkpoint inhibition (ICi). However, the factors determining the level of infiltration of immune cells and tumor-associated macrophages (TAMs) of MBM, remain largely undefined. To dissect the cellular composition of the ecosystems of MBM, we performed high-multiplex in situ transcriptomic profiling using the 10x Genomics Xenium platform across 47 tissue cores from MBM (n=34: multiple, n=22; solitary, n=12), adjacent CNS tissue (n=5), CNS control tissue from non-cancer patients (n=4) as well as primary CNS lymphoma (PCNSL, n=2 patients), compiled on tissue microarrays (TMAs) (Figure 1d, left and center panels, Table S1). Primary lymphomas of the central nervous system (PCNSL) are mainly diffuse large B-cell lymphomas (DLBCLs) confined to the central nervous system (CNS)^38,39^ whichwere included here as a reference for comparison with MBMs.

All samples were subjected to Xenium imaging-based spatial transcriptomics and histological analyses (Figure 1d, right panels). Samples were collected from MBM of different areas within the brain of patients who received BRAFi and/or ICi therapy (Figures 1e, Table S1). The multiple brain metastases originated from a patient diagnosed with stage 1b acral melanoma (Figure S1d, S1e, left). The primary tumor had spread to local lymph nodes, bones, the liver, the lungs, and various intracranial areas in the brain. The brain metastases showed no response to binimetinib/encorafenib. (Figure S1e, right). To capture and account for intratumoral heterogeneity, we obtained multiple biopsies (n = 2–3) from most tumors. Dimension reduction using UMAP shows that the individual cells, whose profiles were analyzed from the tissue punches, can be clearly classified into well-defined molecular subgroups (Figure 1f, Figure S1f). Spearman rank correlation revealed clustering of samples primarily regarding their content of admixed CNS tissue which was evident in tumor samples taken at the tumor boarder area which clustered with CNS control samples (Figure 1g) and further revealed a core set of samples with low content of admixed CNS (Figure S1g). The UMAP representation of marker-classified cell types (Figure 1h) enabled a more detailed characterization of distinct tumor cell subsets, validated NGFR-associated genes like SOX4 and BZW2, and revealed the inverse correlation of NGFR with MET receptor (Figures 1i, S1h).

In general, the extent of immune cell infiltration correlates with tumor response to ICB therapy^40,41^. Tumors with high immune and low stromal scores exhibit the most favorable response, whereas those with high scores for both compartments define an immune-excluded phenotype. Conversely, tumors featuring low scores in both stromal and immune compartments represent an immune-desert (cold) phenotype. We applied the ESTIMATE algorithm^42^ for calculation of immune scores, stroma cell scores and ESTIMATE scores for tumor classification. We observed lowest scores in CNS controls and adjacent CNS tissue as expected and the majority of tumors featured unfavorable high immune and stroma scores (Figure 1j). Since the scores are calculated as aggregates across individual cells and thus the specific cell types that determine the score levels are not clearly identifiable,we mapped the immune scores to individual cells and observed that highest immune scores were evident in T cells (CD8A) and TAMs (Figure 1k, left). However, we also observed tumor cells with elevated immune scores, since the score signature also included markers of antigen presentation by cells in general. Highest antigen presentation scores were observed for TAMs, immune(T) cells and dendritic cells (DCs, Figure S2a). We next applied an additional signature to specifically investigate antigen presentation by tumor cells (TCs). This enabled us to stratify the TCs into high- and low-expressing groups (p<2.22e-16) (Figures 1k, right and S2b). Notably, low marker expression was observed in the BZW2- and SOX4-high clusters, as well as in cells displaying a neural crest-like phenotype. Next, we performed analysis for differentially expressed genes among TCs with high and low expression of antigen presentation markers (Table S3). We observed significant enrichment of the mediators of interferon signaling *IFI44L* and MX1 as well as molecules for antigen presentation HLA-DQA1, HLA-B and antigen transport TAP1 among the top upregulated genes in TCs with high expression of antigen presentation markers (Table S3). In contrast, we observed downregulation of marker of a neural crest-like state RELN, IGFBP3, COL12A1 and SOX11 (Table S3).

### Spatial profiling revealed patterns of immune suppression in MBM

Therapeutic interventions likely foster selection of cellular subsets featuring minimal residual disease (MRD)^43,44^ which escape immune surveillance through downregulation of MHC class I molecules. To further investigate the landscapes of antigen presentation of MBM, we selected a subset of MBM samples which represented different independent patients and treatment strategies (Figure S2c) and featured a high number of transcript counts (Table S1) ensuring robust downstream analyses. Samples with extremely low cell counts (<1,500 counts) were excluded from the final spatial analysis because severely restricted cell numbers undermine statistical power, impair the identification of rare cell states, and compromise the analysis of local cell-cell interactions. Hence, we performed a thorough analysis of a core set of TMA samples for identification of the spatial single cell heterogeneity of MBM across different patients who received BRAFi and/or ICi therapy and to investigate the patterns of immune cell infiltration across multiple progression stages. Unbiased single-cell clustering of the selected TMA core set, combined with automated H&E image alignment (Methods), revealed distinct cellular subclusters defined by key molecular phenotypes (e.g., Ecad⁺, NGFR⁺, MET/SOX4⁺; Figure S3a, b) and cleanly separated tumor cells from CNS tissue and primary CNS lymphoma (PCNSL; Figure 2a, b). Importantly, these results demonstrate that the core set faithfully captures the molecular features and heterogeneity of the entire initial sample cohort.

**Figure 2:**
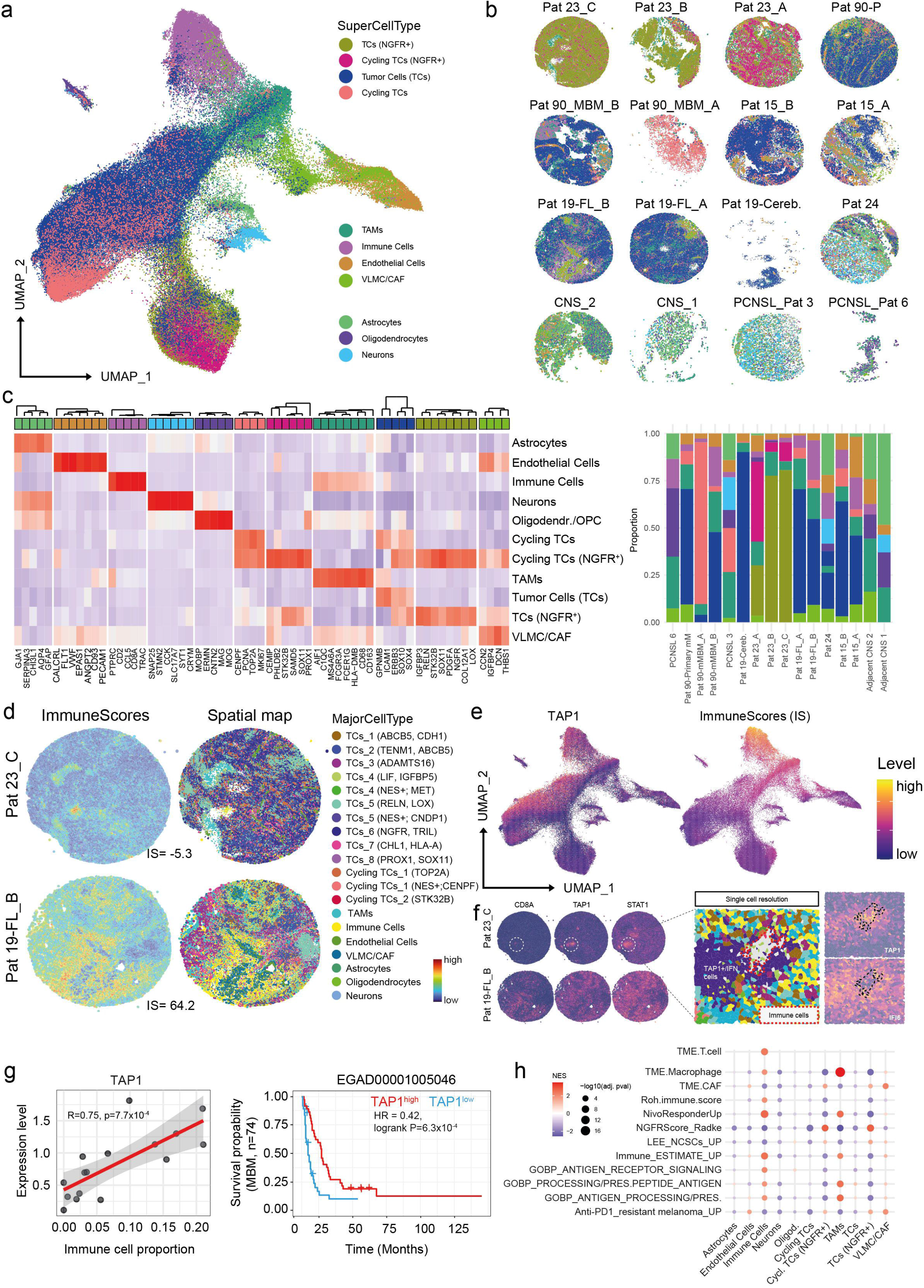
Spatial deconvolution of MBM revealed defined molecular stages. **a.)** UMAP-projection of superior cell types (“SuperCellTypes”) of aggregated tissue cores indicating tumor cell populations and CNS-derived cellular subsets. Cell types are indicated. **b.)** Spatial mapping of SuperCellTypes indicating different stages of tumor heterogeneity and development. **c.)** Left panel: Heat map presentation of top markers of cell types indicated in (a) showing their distinct marker-based classification. Right panel: Proportions of SuperCellTypes in tissue specimen shown in (a, b). **d.)** Left maps: Spatial mapping of immune scores, right maps: mapping of detailed MajorCellTypes indicating marker-defined tumor cell subsets in tissue cores of immune cell-excluded and infiltrated tumors. **e.)** UMAP-projection of levels of TAP1 and immune scores (IS) in single cells of tissue cores, the level is color coded. **f.)** Left maps: Spatial mapping of CD8A, TAP1 and STAT1 expression in tumors shown in (d). Right panels: Single cell resolution of an immunogenic niche in a tumor with an otherwise immune-excluded phenotype (Pat 23C) and mapping of TAP1 and IFI6. **g.)** Left panel: Dot plot of the immune cell proportions across all TMA cores and TAP1 expression level, shows significant correlation (R= 0.75, p=7.710^−4^). **h.)** Enrichment plot showing NES (normalized enrichment scores) of selected signatures across SuperCellTypes.

Cell-type identity was assigned by evaluating canonical marker genes (Table S4), enabling high-resolution annotation of tumor cells (TCs, cycling TCs) and diverse microenvironmental subsets,including TAMs, astrocytes, neurons, and endothelial cells (Figure 2a). Including PCNSL alongside MBM provided a critical comparative framework: despite their distinct developmental origins, both malignancies remodel the brain microenvironment, allowing us to disentangle tumor-intrinsic factors from microenvironmental cues driving progression and immune evasion. Spatial mapping of cell types generated 16 core-specific maps, revealing marked heterogeneity within tumors, between matched cores, and across patients (Figure 2b).

Cellular subgroups were defined based on a set of marker genes with the highest expression levels in a certain single cell cluster (Table S2, log2FC ≥1, p≤0.05), (Figure 2c, left panel). Initially, TCs were classified only into broad categories (SuperCellType): TCs (general tumor cells), cycling TCs (proliferating TCs: TOP2A, PCNA, MKI67, CDK1, CENPF, as well as NGFR-positive and proliferating NGFR-positive cells. Whereas the SuperCellType summarized tumor cell subsets, the more detailed analysis revealed subclassification of tumor cells (MajorCellTypes, Figure S3c). Cell type proportions demonstrated an expected heterogeneity across patients and tumor cores differences in the level of immune cell infiltration (Figure 2c, right panel, Table S5).

Next, we mapped spatial immune scores across tissue cores to evaluate the overall immune landscape (Figure 2d, Figure S3d). This spatial mapping revealed distinct immunogenic hotspots and highlighted pronounced inter- and intra-sample variability in immune cell distribution. Intriguingly, even tumors categorized as immunologically “cold” by the ESTIMATE algorithm (e.g., Patient 23) harbored localized “hot” microenvironments. To resolve the cellular drivers behind these regional discrepancies, we performed high-resolution spatial mapping of the tumor cell compartment (Figure 2d). A neural crest-like phenotype in Patient 23 was prominently highlighted and probably regulated^37^ by NGFR^+^ tumor subsets co-expressing markers of proliferation (MKI67, CENPF; Cycling TCs_2), hypoxia (IGFBP3^45^, LOX^46^, HILPDA^47^; TCs_5), and neural lineage (TRIL, LIFR; TCs_6). This phenotype constituted the primary molecular difference between Patient 23 and Patient 19 (where it was absent) and, in line with previous findings, underlines the mechanism driving immune cell exclusion in Patient 23. In addition to a neural crest-like phenotype, tumor cell antigen presentation is markedly repressed by hypoxia^48,49^, a state closely linked to NGFR expression^50^. In contrast, “hot” tumor cores displayed a fourfold increase in TAM accumulation, which likely accounts for the augmented immune cell recruitment observed in these regions^51^.

Proper antigen presentation requires the expression of TAP1, a transporter located on the endoplasmic reticulum membrane that transfers peptides to MHC class I molecules^52^ and a well-established target of STAT1/interferon signaling. Downregulation or mutation of TAP1 can promote immune escape^53,54^. We assessed the level of TAP1 expression across single cells of all tumor cores. We observed that TAP1 expression separated tumor samples and revealed lowest expression levels in NGFR^+^ cells (Figure 2e, left panel) and low immune scores (Figure 2e, right panel). We investigated the spatial expression of TAP1 together with STAT1 and CD8A in both the “hot” tumor and an “isolated” hot niche identified within the immune “excluded/cold” tumor of Patient 23. Whereas high expression of TAP1 and STAT1 was observed surrounding CD8A-expressing areas in the “hot” tumor (Figure 2f, left panel), co-localization of TAP1, STAT1, and CD8A was evident only within the single-cell–resolved hot niche of the “cold” tumor (Figure 2f, right panel). A marker analysis of the hot niche revealed high levels of interferon-response genes, e.g. IFI6 of areas surrounding the TAP1^+^ immune cell niche. These findings suggest that immune cell infiltration is associated with local activation of the TAP1/STAT1/interferon signaling axis and may depend on adequate TAP1 expression. Hypothesizing that TAP1 expression may serve as a crucial factor for immune cell infiltration of MBM, we determined the immune cell proportions across samples and observed significant correlation (R=0.75, p=7.7×10^−4^) with TAP1 expression in tumor cells and immune cells (Figure 2g, left panel). According to the hypothesis that TAP1 expression mediates tumor cell recognition by CD8A^+^ T cells, patients with TAP1^high^ MBM exhibited higher survival probability than patients with low expression (Figure 2g, right panel). The distinct expression patterns likely connect cellular subsets with specific molecular features. We performed GSEA using defined gene signatures, particularly predicting the capability of antigen processing and presentations. Antigen processing and presentation pathways peaked in immune cells and TAMs, but were markedly depleted in NGFR^+^ tumor cells (Figure 2h).

### BZW2 and TAP1 discriminate immunologic niches in MBM

Immune checkpoint inhibitors (ICi) achieved durable responses and improved survival in advanced-stage melanoma patients^55^. However, only small and asymptomatic brain metastases are susceptible to ICi^15,56^. The formation of spatially defined immunosuppressive areas probably accompanies tumor development and intrinsically determines ICi-resistant areas within larger tumors.

To assess the TAP1 associated genes in MBM on a global level, we performed differential expression (DE) analysis between TAP^high^ and TAP^low^ cancer cells (Figure S4a, b) from single-cell transcriptome data of therapy-naive MBM (study GSE200218^57^). The association between TAP1 and STAT1 expression is also evident in treatment-naive MBM and in a global context (Figure 3a), and DE analysis identified 1,651 genes (adjusted p<0.01). Among the top 20 upregulated genes in TAP1^high^ cells were TAP1, APOL6, PSMB8/PSMB9, STAT1, interferon (IFN)-responsive genes and NLRC5 (Figure 3b, Table S6).

**Figure 3:**
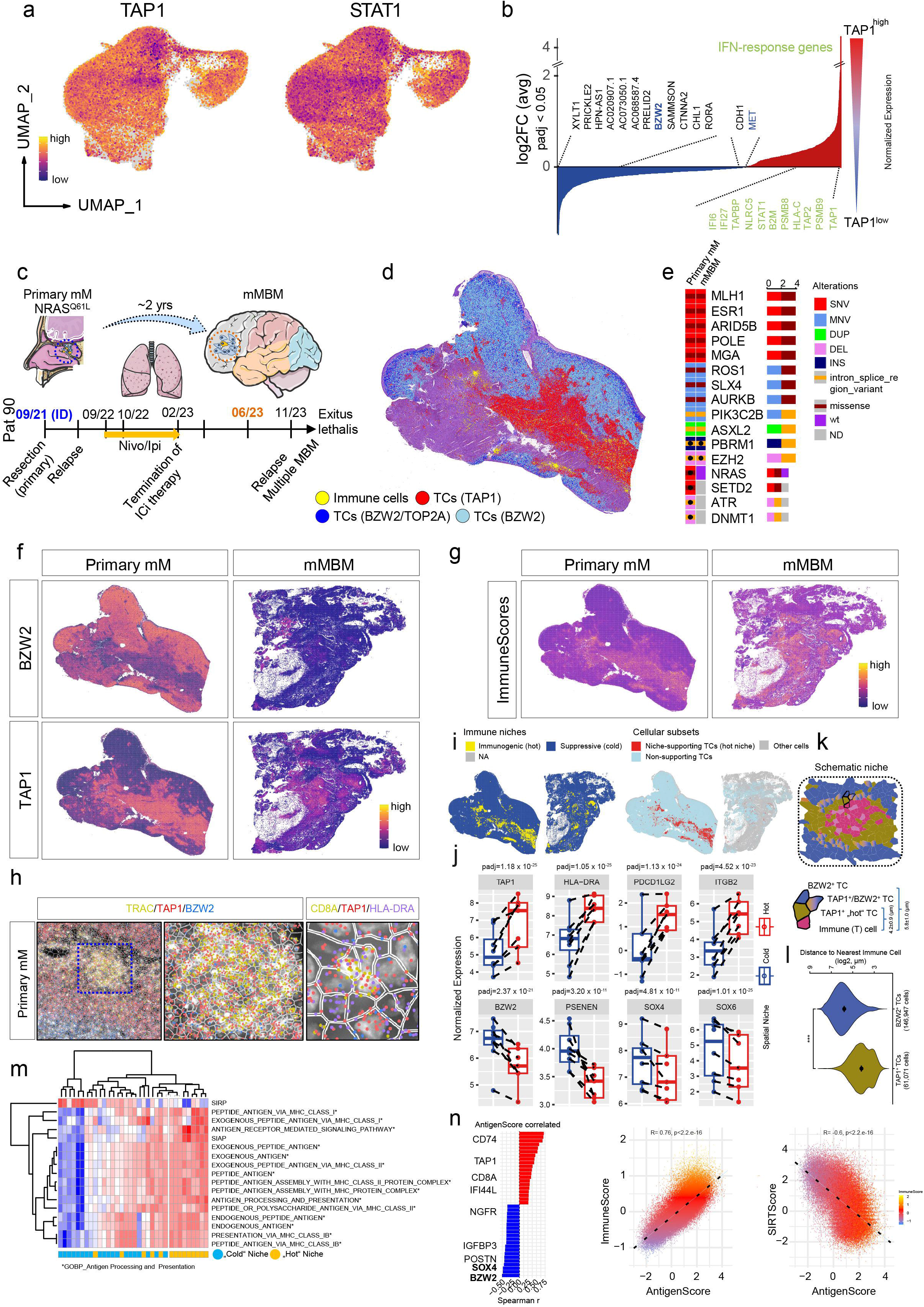
Profiling revealed markers of immune “hot” and “cold” niches. **a.)** UMAP-projection of TAP1 and STAT1 expression across single tumor cells of MBM. Expression levels are color coded. **b.)** DE-analysis of tumor cells of (a) for high or low expression of TAP1. Waterfall plot indicates most expressed (log2FC≥1.05, padj≤0.001) or downregulated ((log2FC≤ −0.9, padj≤0.001) genes. Genes with reported interferon (IFN)-related regulation are highlighted. **c.)** Follow-up scheme indicating the dates of initial diagnosis (ID), treatment regimen and type of metastases emerging. **d.)** Spatial mapping of areas of immune cells (yellow) and tumor cells expressing TAP1 (red), BZW2 (light blue) and cycling BZW2+ cells featuring co-expression of TOP2A (dark blue). **e.)** Presentation of genetic variants retrieved from the profiling of the primary mucosal melanoma (mM) and concordant brain metastasis (mMBM) using the TrueSightOncology (TSO) 500 panel. Variant types and alterations are color coded. SNV = single nucleotide variant, MNV = multiple nucleotide variants, DUP = duplication, DEL = deletion, INS = insertion. **f.)** Spatial mapping of expression levels (color coded) of TAP1 and BZW2 in the primary mM and mMBM showing mutual exclusivity. **g.)** Spatial expression of immuneScores in tumors shown in (f). **h.)** Left and center: Single cell-resolved immunogenic niche, showing molecules of TAP1 (red), BZW2 (blue) and TRAC (yellow, T-cell Receptor Alpha Constant). Right: Molecular presentation of CD8A (yellow), TAP1 (red) and HLA-DRA (purple) in the niche **i.)** Illustration of automatically detected (hot spot analysis) immunogenic “hot” (yellow) and suppressive “cold” (blue) areas within a represented set of tumors. **j.)** Box plots showing levels of selected representative markers for “hot” niches: TAP1, HLA-DRA, PD-L2 (PDCD1LG2) and ITGB2 (upper row) and “cold” niches: BZW2, PSENEN, SOX4 and SOX6 (lower row) as determined by DESeq2 analysis. All markers were significantly differentially expressed between “hot” and “cold” areas across all aggregated niche-building tumor cells of selected aggregated tumor samples (TMA and non-TMA). Box and whisker plots show median (center line), the upper and lower quartiles (the box), and the range of the data (the whiskers), including outliers. **k.)** reconstruction of an immunogenic niche, containing immune (T) cells (pink) and TAMs (light pink) which are surrounded by TAP1^+^ “hot” tumor cells (TCs, greenish brown). The immunogenic niche in turn is embedded in the suppressive area by BZW2^+^ cells (blue) and intermediate cells (color gradient). Lower scheme: enlarged illustration of cells indicated above indicting the distances of TAP1^+^ and BZW2^+^ cells to nearest immune cells. **l.)** Violin plot indicating the distances of immunogenic and suppressive tumor cells to nearest immune cells as calculated across n=61,071 TAP1^+^ and n=146,947 BZW2^+^ TCs. In b, c significance was determined by unpaired t-test, in **(j)** significance was determined by DESeq2 analysis. **m.)** Single samples GSEA of “hot” and “cold” niches/areas for enrichment of indicated GOBP (GeneOntology, BiologicProcesseses) signatures and SIRP (spatial immune-repressive program) and SIAP (spatial immune-activation program). Niches and NES levels are color coded. **n.)** Left panel: Bar plot showing correlation coefficients (r, Spearman rank) of the top20 genes with significant (padj≤0.01) correlation to the AntigenScore. BZW2 and SOX4 (highlighted) show the most negative correlation. Center and right panels: Dot plot showing the positive or negative correlation of AntigenScore and ImmuneScore (R= 0.76, p<2.2×10^−16^) or SIRTScore (R= −0.60, p<2.2×10^−16^) of single cells across all TMA cores. The level of ImmuneScore is color coded in both plots.

Although all genes are essential, NLRC5 serves as master transcriptional regulator of the antigen processing and presentation machinery^58^. Moreover, we identified the neuronal cell adhesion molecule CHL1, SAMMSON (long non-coding RNA), CTNNA2, RORA and BZW2 among the top20 downregulated genes in TAP1^high^ cells, thereby independently validating the inverse relationship between the translation regulator and the antigen-presenting machinery. Moreover, a pan-cancer analysis suggests that BZW2 shapes a non-inflamed, immunosuppressive microenvironment across multiple cancers and inversely correlates with immune checkpoint markers^59^. Comparing individual cells based on BZW2 expression levels demonstrated that BZW2^+^ cells significantly downregulated key antigen presentation components, including LGALS3, HLA-B, HLA-C and TAPBP. Conversely, other marker such as MET, SOX4 and SOX6 were markedly upregulated in this population (Table S6).

Based on the hypothesis that BZW2 may define the spatial immunosuppressive landscape of MBM, and given the limitations of tumor punches, we investigated the spatial distribution of BZW2 in whole tumor sections from an NRAS (NRAS^Q61L^)-mutated primary mucosal melanoma (primary mM) of the nasal cavity. The primary tumor or the recurred primary tumor progressed to the frontotemporal lobe of the brain under ICi therapy and was resected 4 months after ICi-blockade was terminated (Figure 3c). The spatial mapping of CD8A, TAP1 and BZW2 expression revealed clearly defined areas of co-occurrence (TAP1^+^) or exclusion (BZW2^+^) of CD8A T cells (Figure 3d).

Tumors displaying immune exclusion may progress primarily due to a failure of effective immune surveillance. To investigate this, we compared TAP1 and BZW2 expression levels between primary tumors and brain metastases. Apart from the loss of NRAS^Q61L^ in the mMBM, both tumors were genetically identical (Figure 3e) but showed a distinct pattern of expression of BZW2 and TAP1. Surprisingly, we observed increased TAP1 expression accompanied by a loss of BZW2 expression (Figure 3f) and increased immune scores (IS), IS(primary mM) = 21.2 vs. IS(mMBM)=79.7 (Figure 3g). However, TAP1 expression was primarily detected in TAMs rather than in tumor cells. Since tumors have developed in significantly different environments, they likely exhibit distinct ecosystems that are only partially comparable.

Next, we dissected an immune (T) cell niche for unraveling the “hot” and “cold” areas. Even on a very local level we observed a distinct separation of sites showing high expression of TAP1 and BZW2 (Figure 3h, left panel) with TRAC (T Cell Receptor Alpha Constant) expressing T cells or activated T cells expressed CD8A, TAP1, and HLA-DRA located within the TAP1^high^ area (Figure 3h, center and right panel). Moderate to low expression of both TAP1 and BZW2 characterized the transition zone.

To gain deeper insight into the cellular composition of niches, we performed hot spot analysis to detect local microenvironments with IS^high^ cells (see Method section). Hot spot analysis automatically classified each cell as “hot” if the proximity test (Getis-Ord test^33^) for that gene was significant, indicating that the cell is located near a region where IS^high^ cells are abundant; otherwise, it was classified as “cold”. Considering the various stages of development, it is likely that immunogenic (“hot”) and suppressive (“cold”) niches emerge and regress over time during disease progression, as revealed by automated niche mapping (Figure 3i, left panels, and Figure S5a), thereby shaping tumor landscapes and treatment responses. We observed that TAP1^+^ tumor cells were the main contributors of immunogenic niches; however, only a subset of niche-supportive tumor cells were maintained during brain metastasis or during the establishment of the brain tumor (Figure 3i, right panels and Figure S5a). Moreover, we found general (other) markers that did not significantly classified niches. In order to identify valid markers that characterize cold and hot niches independently of the tumor’s mutation and therapeutic status, we performed DE analysis (see Method section) of the previously defined TAP1^+^ tumor cell niches surrounding IS clusters (group of cells with high IS). Differential expression analysis of TMA samples and whole-tumor sections, evaluating tumor cells both in isolation and within integrated tumor-microenvironment niches, validated a core set of immunogenic niche markers such as TAP1, HLA-DRA, PD-L2 (PDCD1LG2) and ITGB2, as well as NKG7, CCL5, CD8A and C1QC (Figure 3j, upper row; Figure S5b; Table S7). In contrast, expression of BZW2, PSENEN, SOX4 and SOX6 as well as MET and IGFBP3 classified the potential immune suppressive/immune cell depleted environment (Figure 3j, lower row, Figure S5b). In addition to single genes, we applied state-predictive gene signatures for calculation of scores (Table S6) and observed significant enrichment of immune-related molecular programs, particularly ESTIMATE, Roh, IPRES (innate PD-1 inhibitor resistance^60^) and NRS (Neoadjuvant Response Signature^18^) in “hot” niches. In addition to immune cells, we observed a significant enrichment of TAMs expressing C1QC, TMIGD3 and AIF1 in immune “hot” niches. The comprehensive niche profiling uncovered differential niche-dependent regulation of additional, immune cell-specifying signatures and revealed NGFR-related programs unrelated to BZW2^+^/SOX4^+^-associated immune suppressive niches. In line with our data, we suggest that immunogenic niches comprise an immune cell/TAM enriched core which is surrounded by TAP1^+^ tumor cells embedded into the BZW2^+^ suppressive environment (Figure 3k, upper scheme). The model is supported by the distance of TAP1^+^ and BZW2^+^ tumor cells to the nearest immune cell, defined by the expression of a gene signature including markers such as CD3G, CD8A, CD4, TRAC and PTPRC. Proximity calculations across all niches revealed average distances of 4.2 ± 0.9 µm for TAP1^+^ cells (n = 61,071) and 5.8 ± 1.0 µm for BZW2^+^ cells (n = 146,947) to the nearest immune cell (Figures 3k, lower scheme and 3l; Table S8), respectively.

The recognition of tumor cells by T cells requires proper antigen presentation. We therefore investigated the antigen presentation patterns of the different cellular subclusters. To this end, we performed a single-sample gene-set enrichment analysis (ssGSEA) using Gene Ontology (GO)/Biological Processes (BP) signatures that specify the mechanism of antigen processing and presentation (Figure 3m).

Moreover, we defined SIRP (spatial immune resistance program) and SIAP (spatial immune activation programs) using the top enriched genes deduced from niche profiling. We found that “hot” niches exhibited low SIRP or high SIAP accumulation. Moreover, we observed enrichment of antigen presenting programs predominantly in the “hot” niches (Figure 3m). With regard to the ability to present antigens, we have defined an AntigenScore that measures not only TAP1 but also the expression of MHC class I and II genes (HLA-A, HLA-B, HLA-C, HLA-DMB, HLA-DPB1, HLA-DQA1, HLA-DRA, TAP1; Table S9). We calculated the score across all TMA cores and defined genes which significantly correlated with the score and validated BZW2 and SOX4 as inversely correlated genes. Moreover, we found positive (R= 0.76, p<2.2×10^−16^) and negative (R= −0.6, p<2.2×10^−16^) correlation of the antigen scores with immune and SIRT scores demonstrated (Figure 3n). In summary, we provided evidence that TAP1, BZW2 and/or SOX4, among other yet uncharacterized markers, define spatial landscapes of immune activation and suppression and may influence the response of MBM to ICi therapy.

### Single cell-resolved spatial profiling uncovered the constitution of immunogenic niches

We hypothesize that the ratio and spatial dimensions of ‘hot’ and ‘cold’ niches govern a tumor’s overall immunogenic phenotype and its sensitivity to immune checkpoint blockade. By aggregating expression profiles of cells within specific niches, we identified signature markers distinguishing “hot” from “cold” intra-tumoral microenvironments. To trace compositional shifts during tumor progression and resolve these immunogenic states spatially, we conducted a comparative marker analysis across tumor sections.

Spatial profiling of ∼410,000 cells from the primary melanoma (mM) and 290,000 cells from the associated brain metastasis (mMBM) revealed a significant change in cellular composition as the tumor progressed. The spatial maps resolved up to 12 distinct tumor cell subtypes (including 4 cycling subsets), alongside TAMs, infiltrating immune cells, vascular-leptomeningeal cells, cancer-associated fibroblasts (VLMCs/CAFs), host epithelial cells, and MBM-associated astrocytes (Figures 4a, b; S6a, b). The loss of mucosal epithelial cells and gain of CNS-related cells such as astrocytes during the process of brain metastasis was expected. Brain metastatic progression markedly altered tumor subtype composition, with specific subsets either emerging or disappearing during metastasis. For example, a rare SLC17A6^+^ (TCs_3) subset, a glutamate transporter-expressing cell type of uncertain prognostic relevance, was present in the primary tumor but virtually absent in the brain metastasis (Figure S6a, b). In contrast, we observed an increase in cellular subsets expressing the MET receptor (TCs_2, Cycling TCs_1; Figure S6a), suggesting a potential role for MET receptor signaling during brain metastasis and its possible influence on immune cell infiltration patterns. Furthermore, we observed enrichment of VLMC/CAFs and immune cells (Figure S6c, left) in line with previous observations^61^.

**Figure 4:**
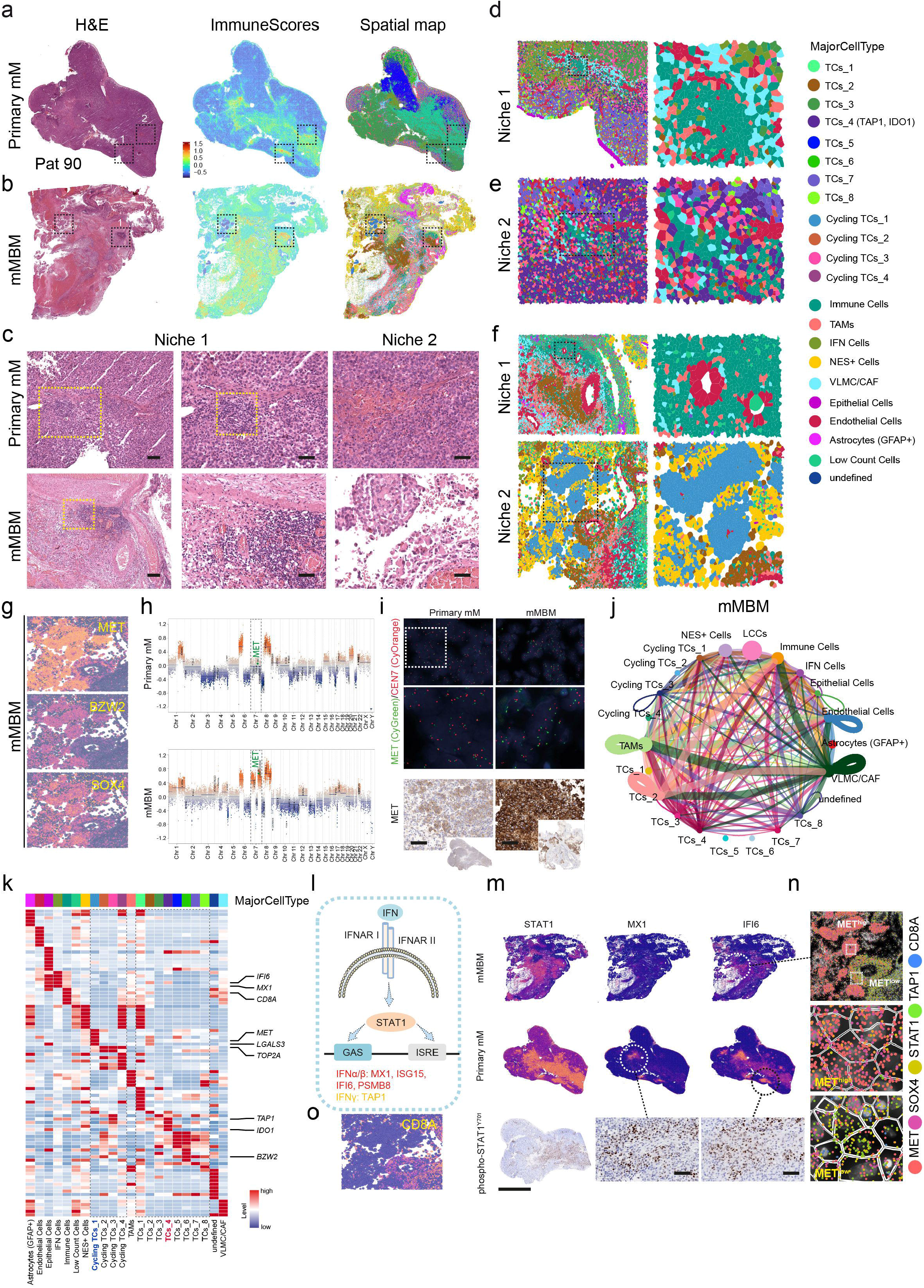
The single cell resolution of niches. a.-b.) Left panels: H&E-based selection of immunogenic niches and illustration of immune scores (center panels) and spatial localization of niches (right panels). **c.)** Magnified areas of immunogenic niches (n=2) of primary mM (upper row) and mMBM (lower row). **d.-e.)** Spatial mapping of MajorCellTypes defined niches showing tumor cell diversity and niche-forming cells of the primary mM. **f.)** Spatial mapping of MajorCellTypes defined niches of the mMBM. MajorCellTypes are color coded. g.) Spatial mapping of expression of MET, BZW2 and SOX4 of an mMBM-specific niche (Niche 2). **h.)** Methylome-based copy number variation (CNV) profiles of the primary melanoma (mM; upper panel) and the matched melanoma brain metastasis (mMBM; lower panel). Chromosome 7 gain resulting in MET amplification in the mMBM is highlighted and compared with the primary mM. **i.)** Left: Fluorescence in situ-hybridization (FISH) analysis (upper panel) and IHC (lower panel) for MET of tumor cells of the primary mM and of mMBM (right). MET(CyGreen) or CEN7(CyOrange) probes were applied. **j.)** Illustration of cell-cell communication of different cellular subsets of mMBM as retrieved from CellChat analyses. Strong communications are indicated by thicker lines, communications within a cell cluster is indicated by loops. **k.)** Heat map illustrating markers of MajorCellTypes of cellular subsets of the primary mM and of mMBM. Selected marker genes are highlighted. **l.)** Simplified scheme of IFN/STAT1 signaling, specific targets are shown. **m.)** Spatial mapping of levels of STAT1, MX1 and iFI6 in mMBM (upper row) and in the primary mE (lower row) indicating a niche-associated distribution of gene expression. Lowest row: IHC showing STAT1 phosphorylation at tyrosin (Y) 701 which is required for STAT1 activation. Circles indicate magnified areas. **n.)** Molecule and single cell resolution of a selected niche (Niche 2) of mMBM indicating the localization of MET, SOX4, TAP1 and CD8A.

Next, we selected morphologically evident niches in the primary mM and the concordant brain mMBM (Pat 90) featuring high immune scores (Figure 4a, b) and accumulated histologically classifiable lymphocytes (Figure 4c). The marker-based classification of selected niches revealed in the primary and brain metastatic tumors identified VLMC/CAFs (DCN/THBS1^+^), TAMs (FCGR1A/CSF1R^+^) and endothelial cells (VWF/CALCRL^+^) as important interaction partners and niche forming cells (Figure 4d, e, S6a; Table S10). Moreover, niche profiling validated the close proximity TAP1^+^ tumor cells (TCs_4), but not tumor cells with high BZW2 expression to immune cells (Figure 4e).

Prompted by the marked elevation of MET, alongside BZW2 and SOX4, specifically in mMBM tumor cells (Figure 4f, g), we conducted genome-wide methylation-based CNV profiling of matched primary mM and mMBM samples. Profiling revealed a gain in MET copy number resulting from chromosome 7 polysomy (Figure 4h), which was confirmed by fluorescence in situ hybridization (FISH) using probes targeting MET (CyGreen) and a centromeric control (CEN7, CyOrange; Figures 4i, upper panels; S6d; Table S11). Elevated MET expression in mMBM was further validated at the protein level via immunohistochemistry (Figures 4i, lower panels). Notably, MET amplification persisted in the relapsed tumor, suggesting that MET amplification and activation may act as a druggable molecular driver of brain metastasis. The niche-dependent maintenance of stem-like cells or immune cells requires cell-cell communication^62^. We assessed the capability of communication of the tumor subsets with environmental cells such as immune cells, TAMs and VLMC/CAFs using CellChat analysis of primary mE and mMBM of the whole sections and characterized the composition of defined niches^63^. Particularly, TAP1^+^ tumor cell subsets of primary mE (TCs_4, niche 2, Figure S6e, left) and mMBM (TCs_2, niche 2) communicated significantly (p<0.01) with immune cells and VLMC/CAFs whereas tumor cells with amplified MET (Cycling TCs_1, niche 2) or expression of BZW2 (TCs_5, TCs_6) were silent (Figure 4j, k). The proposed mechanism of TCs_2 mMBM celĺs communication with CD8A T cells includes HLAs A-C on one hand and APP-CD74 for communication with immune cells and TAMs (Figure S6e, right).

The expression of TAP1 is controlled by interferon signaling (Figure 4l) in dependence of STAT1^64^ activation via phosphorylation at Tyr701, inducing dimerization, nuclear translocation, and DNA binding^65^. We assessed the spatial expression patterns of STAT1 and interferon-stimulated genes MX1 and IFI6^66^. STAT1 was abundantly expressed in both primary mM and mMBM (Figure 4m, first column) whereas MX1 and IFI6 exhibited more site-specific expression patterns. Indeed, MX1 and IFI6 expression was enriched within immunogenic niches (Figure 4m, second and third column). Moreover, phosphorylated STAT1(p-STAT1^Y701^) localized to regions exhibiting MX1 and IFI6 expression in the primary mM, indicating active interferon signaling, a known inducer of TAP1 expression. Investigation of the MET-amplified (MET^high^) niche (mMBM, Niche 2) on the molecule level showed that MET^high^ niches featured significantly low expression of TAP1 (p=0.034), STAT1 (p=0.034) and low level of CD8A^+^ cells (p=0.031). In contrast, we observed enrichment of SOX4 (p=0.0091) and BZW2 (p=0.029) in MET^high^ vs MET^low^ niches, suggesting potential association of SOX4 and BZW2 with MET expression (Figure 4n, Figure S6f). CD8A^+^ T cells were only present in close proximity to TAP1 cells (Figure 4o). The co-occurrence of STAT1, IFI6, TAP1 and CD8A^+^ T cells was also evident in another niche of the mMBM and marker analysis of the niche revealed once more the heterogeneity of immune cell niches.

### Multiplex-immune fluorescence imaging revealed T cell states

TAP1 serves as a crucial player of the antigen presenting machinery ensuring the proper loading onto and presentation by MHC class I molecules. CD8A^+^ T cells in close proximity recognize and bind to the peptide antigens which activates the T cell-mediated tumor cell killing^67^.

To resolve T cell states within immune-hot versus immune-cold/excluded microenvironments, we performed multiplex immunofluorescence imaging of selected core TMA samples featuring high or low immune scores, using the MACSima platform (Figure 5a). T cell activation was evaluated via an antibody panel, recognizing specific surface markers, including the MHC class II receptor HLA-DR, a hallmark of late-stage activation. In contrast, expression of the checkpoint molecule PD-1 may suggest T cell inactivation^68^. The tumor core of the previously analyzed mMBM (Pat90 mMBM_B) shows high expression of MET, BZW2 and SOX4 at immune cell depleted areas. However, at sites of low expression or absence of BZW2 and SOX4 we observed CD8A^+^ T (5.9%) cells and activated CD8A/HLA-DR^+^ T cells (0.4%, Figure 5a). Tumor cells exhibited expression of PD-L1 (0.9%) but higher expression of PD-1 (2.7%) (Figure S7a). In contrast, we observed 11.2% CD8A^+^ T cells in the tissue core of Pat 19 (Pat19_FL_B) but only a minority of T cells featured expression of HLA-DR (1.1%) and co-localization of HLA-DR and CD8A (0.3%, Figure 5b) despite expression of TAP1. Tumor cells exhibited expression of PD-L1 (0.5%) and expression of PD-1 (3.5%) (Figure S7b).

**Figure 5:**
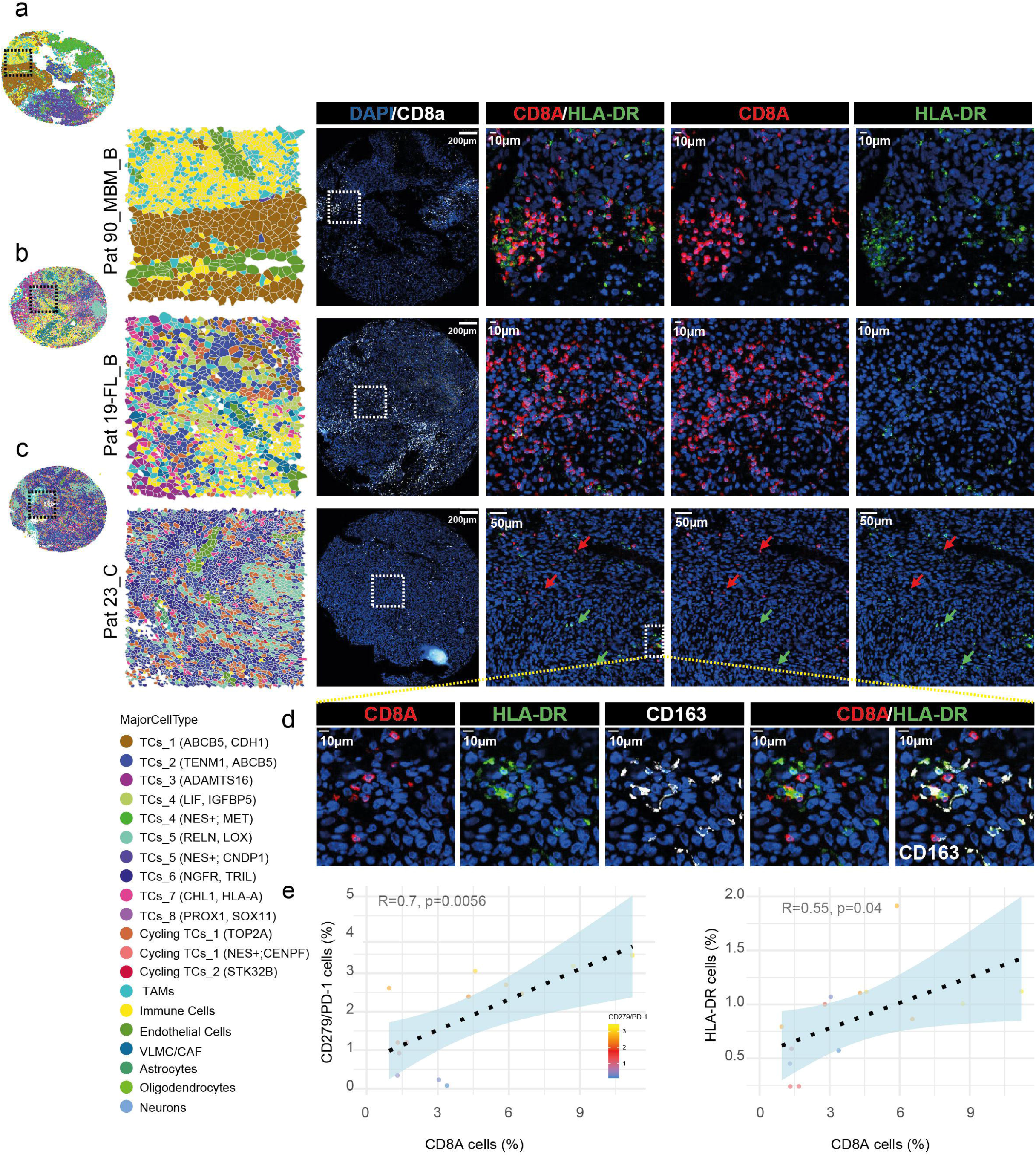
Multiplex-fluorescent MACSima-based imaging revealed heterogeneity of T cell activation. **a.)** From left to right, the region outlined by the black box corresponds to the region outlined by the white box, which is shown at higher magnification in the adjacent images. The magnified view reveals an immune cell-enriched area of the tumor (Pat90_mMBM_B) with immune infiltration and the presence of CD8A+ T cells (red) co-expressing the late activation marker HLA-DR (green) **b.)** From left to right, the black-boxed region corresponds to the white-boxed region, which is subsequently shown at higher magnification. The magnified images reveal an immune cell-enriched region of the tumor (Pat19_FL_B) with immune infiltration at stromal cell-enriched areas (VLMCs/CAFs) and the presence of non-activated CD8A+ T cells. **c.)** From left to right, the black-boxed region corresponds to the white-boxed region, which is subsequently shown at higher magnification. The magnified images highlight an immune cell-enriched region of the tumor (Pat23_C) that largely exhibits immune exclusion, with only sparse CD8A+/HLA-DR+ activated T cells present. **d.)** Investigation of the immune niche shown in (c) reveals T-cell activation and overlapping expression of the inflammatory marker CD163. **e.)** Left panel: Dot plot showing the significant correlation of PD-1 (R= 0.7, p=0.0056 or HLA-DR (R= 0.56, R=0.04) and CD8A cell surface expression.

The tissue core Pat23_C is derived from a highly NGFR, BZW2 and SOX4 expressing, de-differentiated tumor and consequently featured a low immune score. The tumor core contained only 1.4% CD8A^+^ T cells found dispersed throughout the tumor (Figure 5c) contained a minor fraction of 0.04% of activated CD8A/HLA-DR^+^ T cells. Neither PD-L1 (0.3%) nor PD-1 (0.9%) were considerably expressed in the tumor (Figure S7a-c), suggesting that BZW2/SOX4 may serve as alternative mechanisms for suppressing immune cell infiltration and activation. Correlation of CD8A^+^ T cells with HLA-DR and PD-1 revealed two distinct populations suggesting late activated T cells (CD8A/HLA-DR^+^) and potentially exhausted (CD8A/PD-1^+^) cells (Figure 5d, e). In line with the capability of tumor cell elimination by CD8A/HLA-DR^+^ cells, HLA-DRA expression is associated with favored overall survival of MBM patients (Figure S7d).

The polarization of tumor-associated macrophages (TAMs) toward an immunosuppressive, CD163^+^ phenotype represents a critical mechanism driving the phenomenon of immune cell exclusion within the tumor microenvironment (TME)^69^. Determination of CD163 revealed a comparable level of M2 TAMs in all tumor cores (4.2±1.3%; Table S12). In addition to PD-L1/PD-1-mediated inhibition of immune cell infiltration, the presence of M2 TAMs and of FOXP3 regulatory T cells (Tregs) further suppresses T cell activation^70^. Assessment of levels of Tregs in TMA cores, resulted in 0.9±0.8% across all samples with highest proportions in Pat 19 specimen (2.3±0.3%). Considering the different spatial landscapes, CD8A^+^ T cells were present/maintained in all tumors with variable percentages and lowest levels in cores of Pat 23.

### BZW2 and SOX4 serve as promising candidate drivers of immune suppression and ICi therapy resistance

The spatial pattern of lymphocyte infiltration broadly categorizes tumors into immune-inflamed, immune-excluded, and immune-desert phenotypes^71^. Inflamed tumors feature abundant immune cell infiltration throughout the tumor parenchyma, facilitating direct physical contact between CD8A^+^ T cells and tumor cells. In contrast, immune-excluded tumors restrict immune cells to the surrounding stroma, preventing parenchymal infiltration. Immune-desert tumors are largely devoid of immune cells altogether^72^.

Given that spatial profiling linked BZW2 and SOX4 expression to an immune-excluded state, we examined CD3^+^ T cell infiltration patterns across mMBM tissues. CD3^+^ T cells accumulated and infiltrated at sites of TAP1 expression (Figure 6a, left panels) as well as within BZW2^low^ regions (Figure 6a, center panels). In contrast, high BZW2 expression coincided with T cell exclusion (Figure 6a, right panels), indicating that the ratio of TAP1 to BZW2 expression dictates the degree of intra-tumoral T cell infiltration. This finding was independently validated in a clinical cohort of MBM patients (Pts 168, 170) showing complete response (CR) to ICi therapy. We observed a high distribution of CD3^+^ T cells throughout the tumor and low expression of BZW2 (Figure 6b, left and center panels). In contrast, patients (Pat115, exemplary) with progressive disease (PD) showed marginal T cell infiltration and high levels of BZW2 (Figure 6b, right panels).

**Figure 6:**
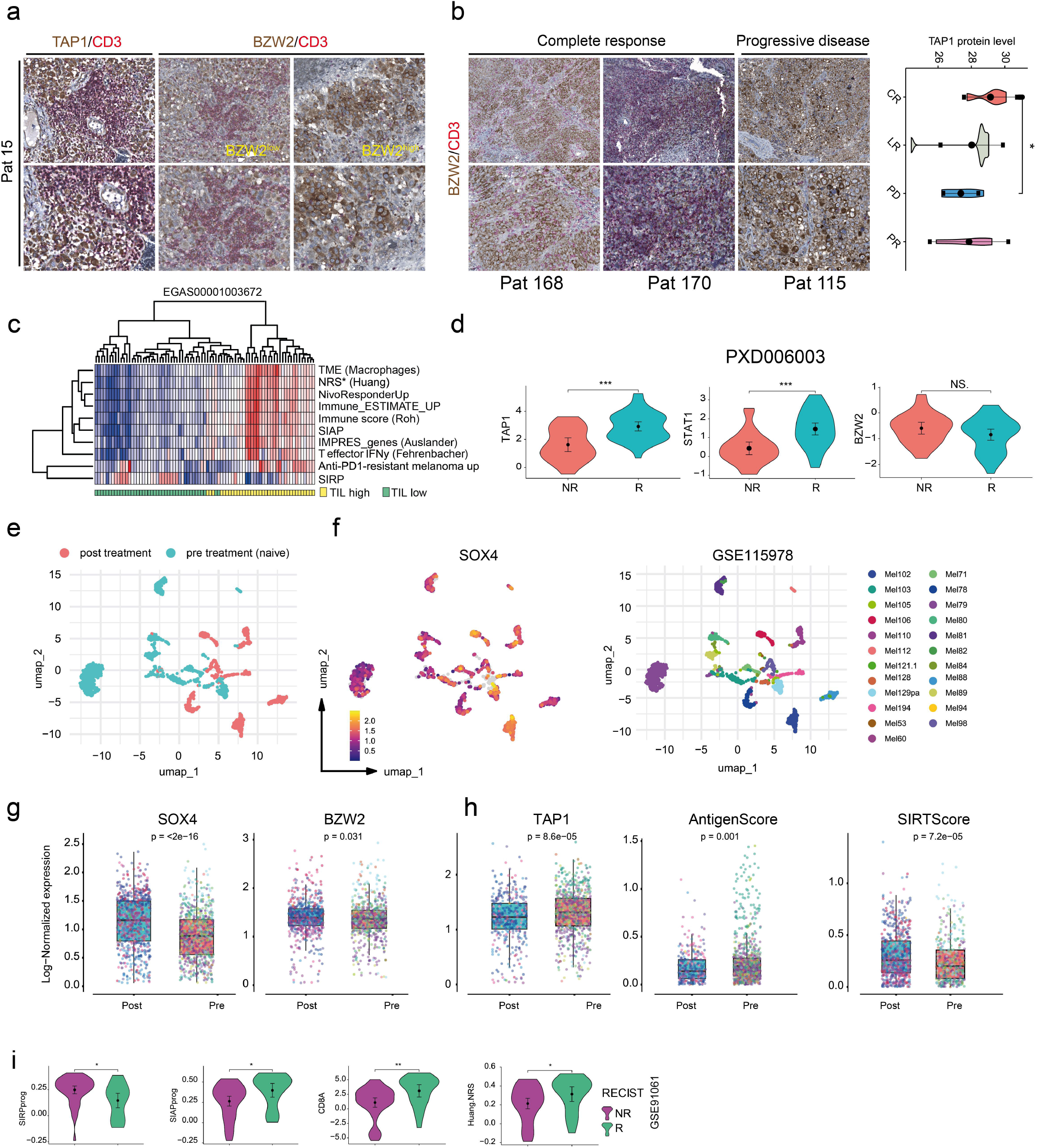
The levels of TAP1, BZW2 and SOX4 determine the response of melanoma to ICi therapy. **a.)** Left panels: IHC for the level and localization of CD3 and TAP1 in a progressive but immune infiltrated tumor, showing CD3^+^ T cells co-localized with TAP1+ tumor cells. Center panels: dispersion of CD3+ T cells at areas of low BZW2 expression in tumor cells. Right panels: absence of CD3+ T cells at areas of high BZW2 expression in tumor cells. **b.)** Left and center panels: presence and distribution of CD3+ T cells at areas of low or absent BZW2 expression in MBM (Pts 168, 170) which showed complete response (CR) to ICi therapy. Right panels: exclusion of T cells in a progressive, non-responding (PD) MBM. Far right: TAP1 protein expression in MBM featuring CR, late relapse (LR), progressive disease (PD) or partial response (PR). **c.)** Separation of MBM (n=79, study EGAS00001003672) regarding the level of infiltration of tumor-infiltrating lymphocytes (TIL) by ssGSEA and gene signatures indicating clinical benefit and favored prognosis: macrophages in the TME or responses to ICi therapy (IMPRES, T-effector-interferon-γ-associated gene expression, neoadjuvant response to anti-PD1 therapy; NRS, Nivolumab response) and prognostic immune signatures (ESTIMATE, Roh) and SIRP and SIAP. **d.)** Violin plots indicating the protein levels (study PXD006003) of TAP1, STAT1 and BZW2 in melanoma with non-response (NR) or response (R) to ICI therapy. **e.-f.)** UMAP-projection of single cells of ICi responding (pretreatment, naïve) and non-responding (post treatment) melanoma (left panel) split by SOX4 expression (center panel) of study GSE115978, which included n=23 patients (right panel). **g.-h.)** Box plots showing expression levels of SOX4, BZW2, TAP1 and levels of AntigenScore and SIRTScore of ICi naïve (pre) or resistant (post) samples tumors. **i.)** Violin plots showing levels of SIRP or SIAP signatures, CD8A or NRS signature (expression of tumor bulk) in patients (n=62) who received ICi therapy and progressed (NR, n=13) or achieved stable disease (R, n=49) in line with RECIST (Response Evaluation Criteria In Solid Tumors) criteria. In a, b scale bars indicate 50 µm. In b, d, g, h, i, significance was determined by t-test.

Proteome analysis of MBM patients showing CR (n=4), PD (n=5) or partial response (PR, n=4) to ICi and late relapse (LR, n=6) revealed significant difference in TAP1 levels between CR and PD patients (Figure 6b, rightmost diagram). Moreover, we evaluated whether SIRP and SIAP signatures indicated the response to therapy. We found that SIRP and SIAP profiles, as well as predictive signatures for response to PD-1 inhibitor therapy, subdivided MBM (n = 79, study EGAS00001003672) based on TIL levels (tumor-infiltrating lymphocytes) (Figure 6c), thereby predicting the immunosuppressed and immunocompetent stages. Furthermore, we assessed the protein levels of TAP1, STAT1 and BZW2 in ICi responder and non-responder (study PXD006003)^73^ showing significantly (p ≤ 0.01) higher expression of TAP1 and STAT1 in ICi responder than in non-responder (Figure 6d, left and center panels). BZW2 was not significantly regulated but showed a trend toward higher levels in non-responder (Figure 6d, right panel). In a comprehensive single cell study (GSE115978, n= 23 patients) Jerby-Arnon *et al.* identified potential mediators of resistance to ICi therapy and identified an immune resistance program which defined the pre- and post-treatment stages (Figure 6e)^14^.

To further evaluate the clinical relevance of our findings, we analyzed SOX4, BZW2, and TAP1 expression levels in ICI-naïve (pre-treatment) and post-treatment/relapsed melanoma patients at both the single-cell and bulk tissue levels. We observed significantly higher levels of SOX4 (p < 2 x 10^−16^) and BZW2 (p = 0.031) in post-treatment/relapsed tumors, underpinning their potential role as mediators of immune resistance by limiting immune infiltration (Figure 6f, g). Strikingly, levels of TAP1 (p = 8.6 x 10^−5^), AntigenScore (p = 0.001) and SIRTScore (p = 7.2 x 10^−5^) significantly distinguished pre- and post-treatment tumors (Figure 6h). Finally, we evaluated the expression of SIRP and SIAP programs, CD8A, and the neoantigen response signature (NRS) in melanoma patients before and during nivolumab therapy^74^. The SIRP program was significantly enriched (p≤0.05) in non-responders (NR), whereas responders exhibited elevated levels of the SIAP program, CD8A, and the NRS (Figure 6i).

Ultimately, spatial expression of BZW2/SOX4 versus TAP1 demarcates immune-excluded from immune-competent regions, with overall ICi response determined by the balance and scale of these opposing niches.

### Phenotypic Heterogeneity Dictates Therapeutic Response in Melanoma Brain Metastases

Concordant, metachronous MBM arising at distinct intracranial sites likely harbor divergent microenvironmental ecosystems, leading to variable therapeutic responses. To unravel the spatial landscapes of concordant MBM from different intracranial sites, we investigated whole-tumor sections of cerebellar (Pat19_Cereb) and frontal lobe (Pat19_FL) metastases (Figure 7a). The tumors showed distinctly different histology, and the mapping of marker-defined cell types revealed remarkable differences. In particular, the frontal lobe tumor exhibited high TAM and T-cell infiltration, which was reflected in the immune score mapping and likely driven by distinct intracranial microenvironments (Figure 7b, c). This pattern was consistent across tumor cores, indicating that relative immune cell proportions served as a key discriminating factor between tumors (Figure 7d). In particular, the frontal lobe tumor exhibited high TAM and T-cell infiltration, which was reflected in the immune score mapping and likely driven by distinct intracranial microenvironments (Figure 7b, c). This pattern was consistent across tumor cores, indicating that relative immune cell proportions served as a key discriminating factor between tumors (Figure 7d). CellChat analysis provides insights into the strength and type of cellular interactions. We observed homogeneity of communications networks, despite different compositions of the cellular landscapes (Figure 7e, f). We identified seven different tumor cell subsets (Figure 7g) and DEG analysis revealed IGFBP5 and HLA-C or SOX4 and ABCB5 among the top expressed genes in the frontal lobe or cerebellar tumor, respectively

**Figure 7:**
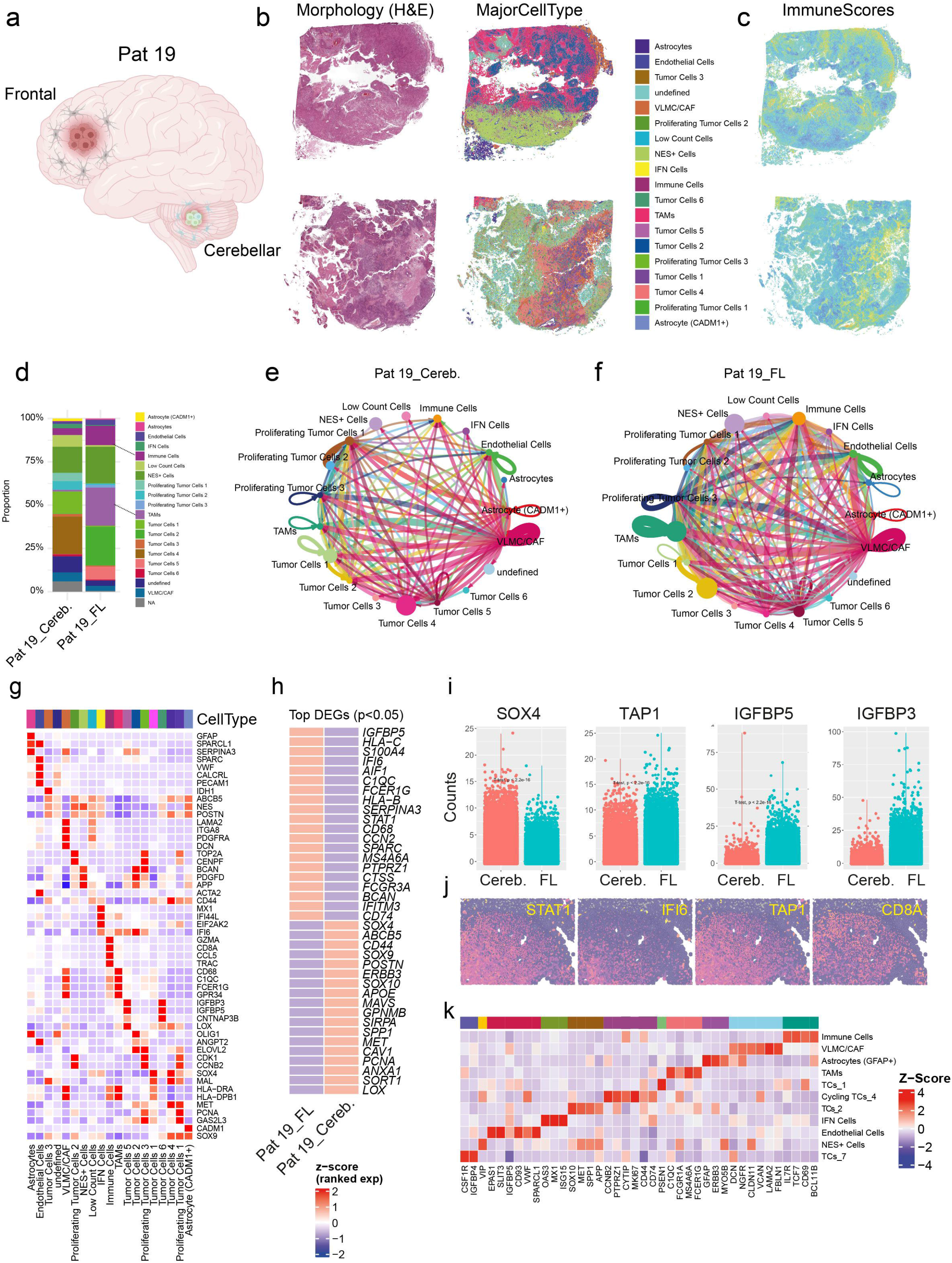
Spatial profiling of concordant MBM from different intracranial sites revealed distinct patterns of progression. **a.)** Scheme showing the intracranial locations of Patient 19 tumors (frontal lobe, FL) and cerebellum (Cereb.). **b.)** H&E staining and spatial map indicating the “MajorCellTypes” of the FL tumor (first row) and of the cereb. Tumor (lower row). **c.)** Mapping of immune scores to tumors shown in (b). **d.)** Stacked bar charts showing the proportions of cell types indicated in (b) of FL and Cereb. tumors. **e.-f.)** CellChat-based analysis of cellular networks in tumors of (b), the strength of communication is indicated by line thickness, the representation of cell types is indicated by the size of filled circles and loops indicate autocrine/self-regulatory processes of a certain cell type. **g.)** Heat maps indicates the top expressed genes by each cellular subset of Patient 19 tumors. **h.)** Heat maps shows the most significantly deregulated genes among FL and Cereb. tumors. **i.)** Violin plots indicate level of expression of SOX4, TAP1, IGFBP5 and IGFBP3 in tumors of (b). **j.)** Spatial mapping of levels of STAT1, IFI6, TAP1 and CD8A to an T cell and TAM-enriched areas of the FL tumor. k.) Heat map representing markers of the area shown in (j).

(Figure 7h). Consistent with our previous observations, TAP1, STAT1, and IFI6 were significantly increased in the frontal lobe tumor, potentially contributing to enhanced T cell infiltration. (Figure 7i, j). Marker analysis of the T cell– and TAM-enriched tumor area identified a core panel of T cell markers: IL7, TCF7, BCL11B, and CD69, suggesting the presence of tissue-resident memory (TRM) T cells, which are associated with a favorable prognosis^75^.

In summary, our findings reveal why concordant melanoma brain metastases (MBMs) show variable responses to immune checkpoint inhibitor (ICi) therapy: elevated interferon signaling and TAP1 expression correlate with favorable outcomes, whereas increased SOX4 and/or BZW2 expression is associated with poor prognosis and immune exclusion.

## Discussion

Tumor ecosystems evolve over time, either by acquiring intrinsic resistance to immune checkpoint inhibitor (ICI) therapy or by developing adaptive immune resistance. Furthermore, as primary tumors metastasize to distant organs, their cellular composition shifts. The brain presents a unique microenvironment; consequently, melanoma brain metastases (MBMs) form distinct ecosystems that differ significantly from primary tumors and demand tailored therapeutic strategies.

In this study, we present an interactive, high-resolution spatial transcriptomic resource of melanoma brain metastases (MBM) and tumor-adjacent CNS tissues. By leveraging 10x Genomics Xenium in situ profiling across 16 core punches and 4 whole-slide sections—including matched primary-metastatic pairs, this publicly available dataset provides a spatially resolved, single-cell map of the MBM microenvironment. Through TissuuMaps, we make this high-multiplex spatial landscape fully accessible to the research community to facilitate further exploration of neuro-oncological niches.

Our approach not only refined previously described molecular tumor cell states, such as the NGFR^+^ phenotype which was clearly associated with expression of IGFBP3, LOX or reelin (RELN) and/or SOX9 expression and proliferative cell states associated with expression of cell cycle genes such as TOP2A, PCNA, MKI67, CENPF and CDK1. Our approach uncovered different states of MET expression in MBM. The MET receptor serves as a promising therapeutic target not only for the treatment of brain metastases in lung cancer patients but also in MBM. Previously, we demonstrated that the MET receptor is amplified as a consequence of chromosome 7 polysomy^35^. Since BRAF and MET are located on the same chromosome, synchronous targeting of both might represent a plausible strategy not only for MBM with acquired resistance to BRAF and immune checkpoint inhibitors (BRAFi/ICi), but also for controlling melanomas that are either BRAF wild-type or harbor non-V600 BRAF mutations. We found MET amplification in a brain metastasis of an NRAS-mutated mucosal melanoma and observed increased MET protein expression in the relapsed primary tumor. Moreover, we observed a loss of the NRAS mutation, which likely occurred in concert with the gain of MET expression. Indeed, the loss of the oncogenic NRAS Q61 mutation alongside MET amplification is consistent with established mechanisms of pathway bypass and clonal evolution in melanoma^76,77^. MET amplification represents a distinct molecular alteration that can sustain downstream MAPK/PI3K signaling independently of primary RAS mutations, thereby driving therapeutic resistance and tumor progression^78^.

MET receptor expressing cells are often correlated with BZW2 or SOX4 and may mediate survival and growth signals and establish certain vulnerabilities. Analysis of MET^high^ niches in the brain metastasis of the primary mucosal melanoma suggested co-expression of MET, SOX4 and BZW2 and low expression of TAP1 and low levels of infiltration of CD8A T cells. However, the functional connection of MET, SOX4 and BZW2 remains elusive, suggesting that neither MET nor NGFR^+^ may regulate BZW2. Indeed, BZW2 and SOX4 were among the top downregulated genes in melanoma cells with knockdown of NGFR^20^.

The BZW2/SOX4/TAP1 axis appears to represent an overarching mechanism operating independently of the E-cadherin/NGFR phenotype. Rather than exhibiting mutual exclusivity, BZW2 and TAP1 displayed a gradual expression continuum across tissue sections. Mechanistically, the upstream regulation of this axis remains to be fully elucidated. Although SOX4 acts as a mediator of TGF-induced epithelial–mesenchymal transition (EMT) in breast cancer^79,80^, this spatial link was not prominent in our cohort. BZW2, a competitive inhibitor of eIF5 that suppresses translation initiation at non-canonical start codons^81^, has been linked to mTOR signaling and a pro-tumorigenic phenotype across multiple malignancies^59^. Conversely, the expression of TAP1 and associated MHC class I antigen-presentation machinery^54^ depends on interferon signaling^64^ and the key transcriptional coactivator NLRC5^58,82^, which is frequently silenced via promoter methylation. Spatial hotspot analysis of immune cell infiltration successfully differentiated immunologically “hot” and “cold” niches. “Hot” niches were defined by a signature of TAP1^+^ tumor cells, HLA-DRA and infiltrating immune populations (including CCL4^+^, ITGB2^+^ and LGALS9^+^ immune cells as well as C1QC^+^, FCER1G^+^ and AIF1^+^ TAMs). To quantify these microenvironmental states, we derived scores for a Spatial Immune Activating Program (SIAP) and a Spatial Immune Suppressive Program (SIRP). Crucially, high SIAP scores predominated in tumors with strong immune infiltration and favorable response to ICi therapy, whereas SIRP expression correlated with non-responsiveness. Predictors of ICi response include a 10-gene IFN\gamma signature containing STAT1 and HLA-DRA. In alignment, “hot” niches coincided with IFN^+^ cells enriched for downstream interferon mediators such as MX1 and IFI6 while STAT1 and HLA-DR(A) proteins localized to areas with low BZW2/SOX4 expression. Although the exact triggers of BZW2 upregulation require further study, we hypothesize that BZW2^+^ cells harbor aberrant STAT1 signaling, impairing TAP1 expression, antigen presentation, and subsequent T-cell recruitment.

In summary, we identify BZW2 and TAP1 as inversely correlated tumor markers that map to distinct spatial niches corresponding to high or low immune infiltration. Operating alongside the established PD-1/PD-L1 axis, the BZW2/TAP1 system represents a novel layer of spatial immune regulation in melanoma brain metastases.

## Declarations

### Consent for publication

“Not applicable”

### Availability of data and materials

A TissUUmaps-based interactive application of annotated Xenium cells and aligned post-Xenium H&E images of all MBM samples are publicly available at https://brain-melanoma-tissuumaps.mdc-berlin.de/. All spatial transcriptomics raw data are deposited at GEO (Gene Expression Omnibus; study GSE306955). Scans of immunohistochemically stained slides were deposited at Zenodo https://doi.org/10.5281/zenodo.16926050. For integrative analyses our study included bulk expression data of MBM (study EGAS00001005975 (https://zenodo.org/record/7013097) and single cell expression data of candidates BZW2, SOX4 and PD1/PD-L1 of melanoma showing response or resistance to immune checkpoint inhibition (study GSE115978); https://portals.broadinstitute.org/single_cell/study/melanoma-immunotherapy-resistance) and therapy-naïve MBM (study GSE200218).

### Code Availability

The R scripts used to analyze TMA and MBM samples as well as scripts to generate the figures are available at https://github.com/LandthalerLab/MBM_Analysis_Manukyan.

### Authors’ contributions

AM, KP, JR, and TR conceived and designed the study, performed experiments, collected data, analyzed the data, and drafted and wrote the manuscript. EW, HR, TC, JA, IP, AA, FR, AL, KK, KJ, and ML performed experiments and provided essential resources. EW contributed substantially to the conceptual development and critical revision of the manuscript. All authors critically revised the manuscript for intellectual content and approved the final version for submission.

## Acknowledgements

We would like to thank all collaborators, technical staff, and laboratory personnel who contributed to this study. We are especially grateful to Michele Peters, Ayad W.H. Dawood, Andreas Wendt, Dana Ristau, Katrin Müller, Tanja Propp, Betty Nedow, Jenny Hagemann, and Diana Krüger for their support with sample preparation, immunohistochemistry, sequencing, analysis, and technical assistance.

## Supplementary figure legends

**Figure S1: Decoding the molecular subtypes of MBM.** a.) IHC for Ecad (upper panel) and MLANA (lower panel) of Patient 24, indicating the presence of Ecad in melanocytic tumors. b.) Circle plots showing the association of phenotype (amelanotic, NGFR^+^; melanotic, Ecad^+^, *color-coded*) with molecular programs and associated genes; log2FC values are shown). c.) Bar chart showing normalized enrichment scores (NES) of gene set-enrichment analyses of MBM subgroups Ecad^high^ and Ecad^low^ and indicated gene signatures. d.) Schematic summary of development of multiple MBM and meningeosis melanomatosa from stage IB acral melanoma. Additional metastases in bone, lymph nodes, lung and liver are indicated (left). Schematic representation of tumor locations (right). e.) MRI of multiple MBM of the patient described in (d) and brain section with tumors (inlay) (left) and legend of tumor locations shown in (d). Summary of the therapeutic sequence applied to mMBM (right). f.-g.) Legend showing the clustered samples and detail image showing clustering of samples with low CNS but high tumor cell content (core TMA set). h.) UMAP presentation of expression of CDH1/Ecad, NGFR, single cell

**Figure S2: The levels of antigen presentation define cellular subsets.** a.) Violin plots, indicating the levels of antigen presentation score across different cell types of MBM. b.) Signature scores in single cells, showing the capability of antigen processing and presentation. c.) Table indicating the patients characteristics of the core-TMA samples.

**Figure S3: UMAP-based clustering revealed molecular subsets.** a.) UMAP of the unintegrated Xenium cells across all TMA cores, illustrating clustering of samples and common and unique cellular subsets. Molecular subgroups are highlighted. b.) UMAP visualization illustrates the clustering of integrated TMA cores according to donor characteristics. c.) Spatial mapping of MajorCellTypes of TMA cores (TMA1.0). d.) Spatial mapping of immune scores of TMA cores (core-TMA).

**Figure S4: Investigation of therapy-naïve MBM (study GSE200218) revealed TAP1 and BZW2-associated genes.** a.) UMAP-projection of single cells of MBM comprising tumor cells, immune cells and CNS-related cells. b.) UMAP-projection of tumor cells of MBM.

**Figure S5: Profiling revealed characteristics of immunogenic “hot” and suppressive “cold” niches.** a.) Single spatial mapping of BZW2 expression of immune scores (first and second row) and composite visualization of BZW2 and immune scores (third row). Automated detection and assignment of immune niches using hot spot analysis (fourth row) and of niche-supporting “hot” tumor cells (fifth row). Expression levels and cellular subsets are color coded. b.) Box plot representation of expression levels calculated across all niche-encompassing tumor and TME cells of selected aggregated tumor samples (TMA and non-TMA). Immune cell markers NKG7 (natural killer cells), CCL5, CD8A (both T cells), C1QC (TAMs) were significantly enriched in “hot” niches. Levels of tumor cell markers MET and IGFBP3 were calculated across all niche-building aggregated tumor cells. Box and whisker plots show median (center line), the upper and lower quartiles (the box), and the range of the data (the whiskers), including outliers. In b significance was determined by significance was determined by DESeq2 analysis, corrected for multiple testing.

**Figure S6: CellChat identified cell-cell communications of cellular subsets of primary and brain metastatic mucosal melanoma.** a.-b.) UMAP-projection of single cell subsets of primary mucosal melanoma and mMBM revealed compositional changes accompanying metastatic progression. c.) Left: stacked bar chart indicating the proportions of cell types of tumors. Right: levels of BZW2 and TAP1 in tumors shown in (a). d.) Box plots showing signal counts of MET and CEN7 probes. e.) Left: Illustration of cell-cell communication of different cellular subsets of primary mucosal melanoma as retrieved from CellChat analyses. Strong communications are indicated by thicker lines, communications within a cell cluster are indicated by loops. Right: prediction of interaction partners of the cellular subset TCs_2 of mMBM with immune cells and TAMs, the strength of interaction is color coded. f.) Box plots showing transcript counts of a region of high or low MET expression of mMBM (Pat90. Transcript counts of n=4 selected areas within the regions (METhigh/low) are shown. Significance was determined by t-test.

**Figure S7: MACSima analysis revealed low levels of PD-1/PD-L1 in MBM.** a.-c.) MACSima-based multiplex imaging for Tregs (FOXP3), M2 TAMs (CD163) and immune checkpoint markers PD-1 and PD-L1 was performed. d.) Overall survival of MBM patients indicating favorable outcomes associated with high HLA-DRA expression.

---

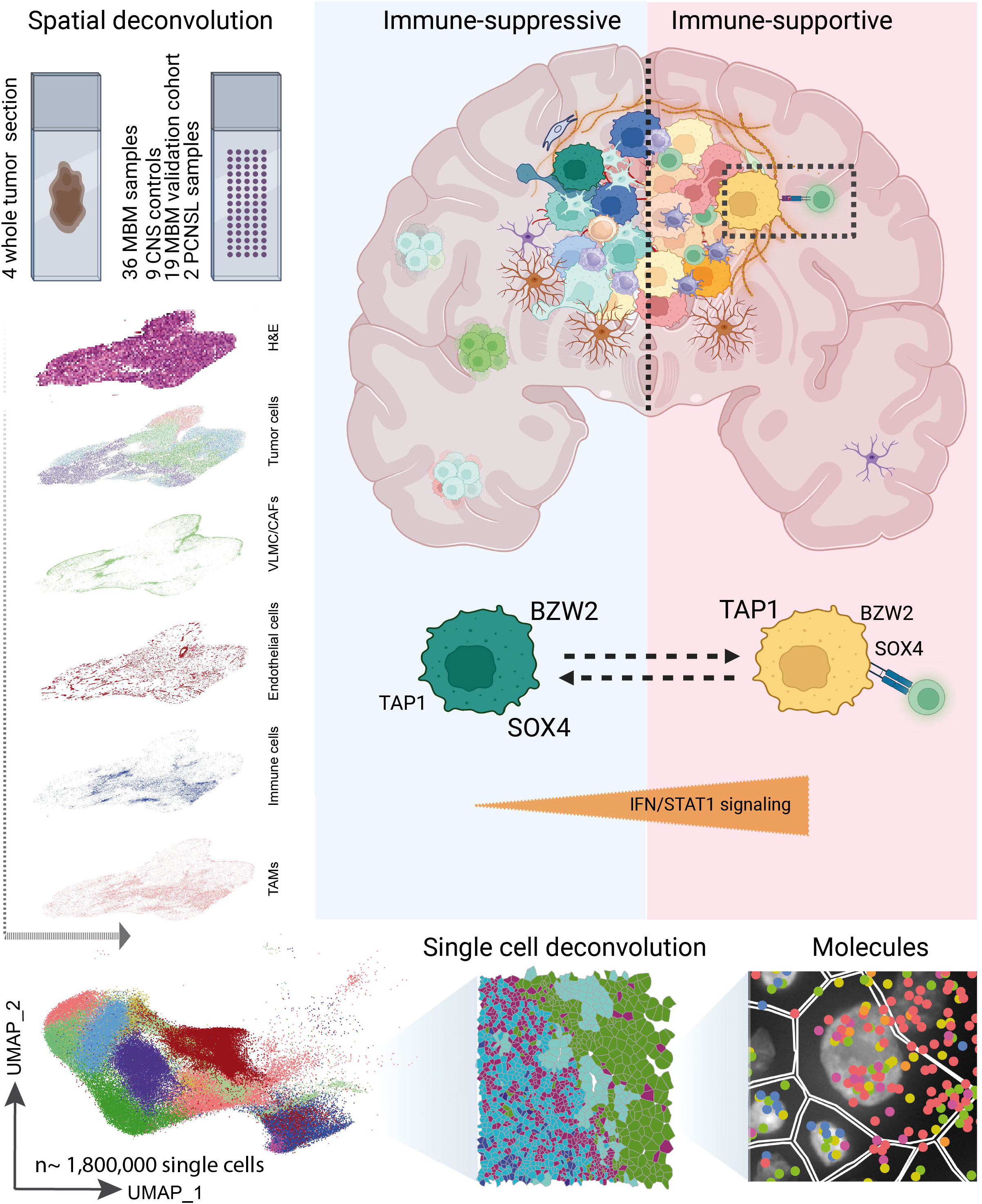

